# Plasma GDF15 affects long-term dementia risk and alters neuro-immune signaling

**DOI:** 10.1101/2025.07.18.25331297

**Authors:** Cassandra O. Blew, Michael R. Duggan, Dimitrios Tsitsipatis, Gabriela T. Gomez, Zulema Rodriguez Hernandez, Luke C. Pilling, Jingsha Chen, Eva Jacobsen, Heather E. Dark, Yifei Lu, Shannon M. Drouin, Cassandra M. Joynes, Minhao Yao, Murat Bilgel, Abhay Moghekar, Qu Tian, Julián Candia, Mary Kaileh, Aditi Gupta, Krystyna Mazan-Mamczarz, Myriam Gorospe, Alexey Lyashkov, Yevgeniya Lukyanenko, Mika Kivimaki, Philipp Frank, Lori L. Jennings, Valborg Gudmundsdottir, Vilmundur Gudnason, Lenore J. Launer, Naoto Kaneko, Shintaro Kato, Makio Furuichi, Masaki Shibayama, Masahisa Katsuno, Keita Hiraga, Yukiko Nishita, Rei Otsuka, James R. Pike, Mary R. Rooney, Pascal Schlosser, Yuhan Cui, Guray Erus, Christos Davatzikos, Rebecca F. Gottesman, Iwao Waga, Priya Palta, Christie Ballantyne, Michael Griswold, Zhonghua Liu, Luigi Ferrucci, Allison B. Herman, Keenan A. Walker

## Abstract

Growth/differentiation factor-15 (GDF15) is a secreted peptide hormone and cytokine that is strongly associated with dementia risk. However, the extent to which plasma GDF15 represents a biomarker and driver of dementia risk remains unclear. Across multiple cohorts, we demonstrated that plasma GDF15 is associated with greater dementia risk over 15-to 25-year follow-up periods when measured in midlife, with stronger associations observed for vascular dementia compared to Alzheimer’s disease (AD). Two-sample Mendelian randomization supported plasma GDF15’s mechanistic role in AD and related dementias, while cohort studies linked it to cerebral small vessel disease, diffuse neurodegeneration, phosphorylated tau, and a cerebrospinal fluid proteomic signature indicative of neuro-immune activation. Exposure of cultured myeloid cells to recombinant GDF15 altered biological pathways that we subsequently demonstrated are predictive of dementia risk, including interferon/antiviral responses, pyruvate metabolism, and scavenging of heme. These findings support circulating GDF15’s role as an early biomarker – particularly for vascular dementia and neuroinflammation – and identify the mechanisms by which it may drive dementia risk.

---

Pathological changes underlying neurodegeneration and dementia can begin decades before clinical symptoms emerge. For example, amyloid-beta (Aβ) accumulation begins for many individuals during midlife, preceding the onset of cognitive deficits in Alzheimer’s disease (AD) by twenty to thirty years^1^. A recent proteome-wide association study^2^ identified dysregulation of immune, metabolic, proteostasis, and synaptic proteins in the plasma of middle-aged adults as early as 20 years prior to dementia onset. Growth/differentiation factor 15 (GDF15)—also known as macrophage inhibitory cytokine-1 (MIC-1) due to its initial classification as an immunosuppressant^3^—showed the strongest association with dementia risk, and was associated with accelerated 20-year cognitive decline and YKL-40 levels (a marker of neuroinflammation) in cerebrospinal fluid (CSF). Together, these findings suggest that GDF15 may function as an early biomarker for dementia and cognitive decline, as well as an indicator of dysregulated immune processes.

GDF15 is a secreted peptide hormone and cytokine of the transforming growth factor-β superfamily^4^ that is expressed primarily outside the central nervous system (CNS; e.g., kidney). Along with being a marker of biological aging^5^, GDF15 has been implicated in a broad range of biological processes, including immune/inflammatory responses^6,7^, energy homeostasis and appetite^8^, and tumorigenesis^9^. Elevated circulating levels of GDF15 have been reported in a variety of age-related conditions, ranging from all-cause mortality to end-stage kidney disease^10,11^. Some, but not all, studies have demonstrated that higher circulating GDF15 levels are associated with long term dementia risk as well as faster rates of cognitive decline^12,13,14^. While these results indicate that plasma GDF15 may function as an early marker of dementia risk, further investigation is required to (i) determine its predictive value across dementia subtypes and population subgroups, (ii) identify the mechanisms that account for GDF15’s relationship with long-term dementia risk, and (iii) understand how these early molecular changes might be therapeutically targeted to provide further benefits to patients.

Using high-throughput proteomic data from six cohort studies, we examined the association of plasma GDF15 with all-cause and etiology-specific dementia risk, characterized GDF15’s relationships with dementia-related endophenotypes, and identified the biological processes through which circulating GDF15 may contribute to cognitive decline and dementia (**Figure 1**). After replicating the associations of midlife and late-life plasma GDF15 abundance with increased dementia risk and detecting particularly robust associations with vascular dementia (VaD), we found support for plasma GDF15’s mechanistic role in Alzheimer’s disease and related dementias (ADRD) using two-sample Mendelian randomization (MR).

**Figure 1.**
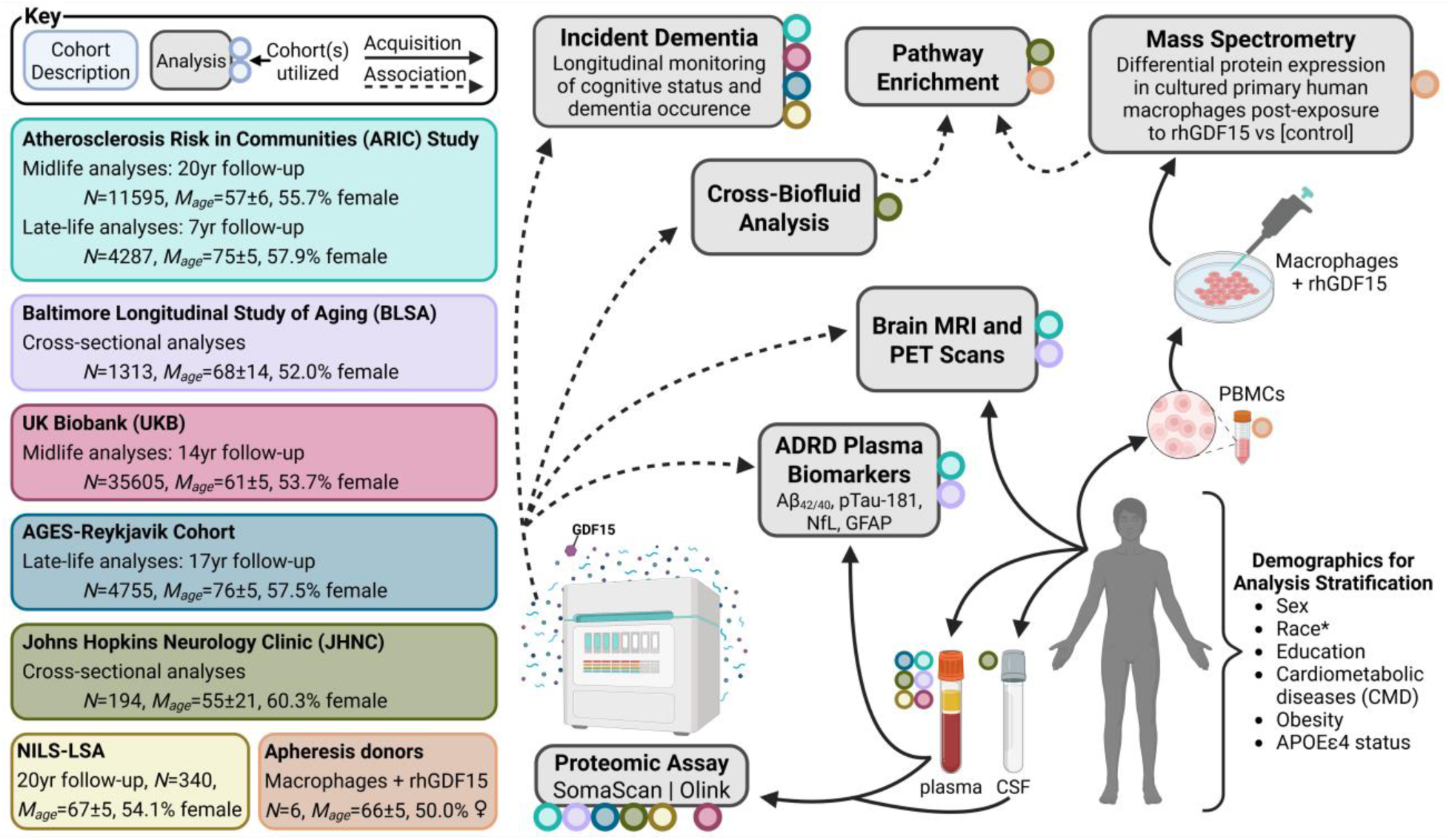
Study design A multicohort approach was implemented to assess plasma GDF15’s predictive value for, and biological links with, dementia risk and dementia-related endophenotypes. Plasma abundance of GDF15 measured on high-throughput proteomic platforms (SomaScan, Olink) was associated with long-term incident dementia across three independent cohorts, as well as neuroimaging and plasma biomarkers across two independent cohorts. The GDF15-associated CSF proteome and the proteome of human macrophages following *in vitro* GDF15 experiments were used for pathway analyses. *Abbreviations*: ADRD, Alzheimer’s disease and related dementias; NILS-LSA, National Institute for Longevity Sciences – Longitudinal Study of Aging; PBMCs, peripheral blood mononuclear cells; rhGDF15, recombinant-human GDF15 *Race-stratified analyses were only conducted in the ARIC study due to race heterogeneity among non-white individuals in other cohorts.

Consistent with these findings, we demonstrated that plasma GDF15 was associated with greater cerebral small vessel disease, diffuse neurodegeneration, phosphorylated tau levels in both CSF and plasma, and a CSF proteomic signature indicative of neuro-immune activation. Subsequently, we exposed cultured human macrophages (differentiated from monocytes isolated from peripheral blood mononuclear cells (PBMCs)) to recombinant GDF15, from which we then identified dysregulated immune and metabolic protein networks that partially mediated the associations with dementia risk. Our results suggest plasma GDF15 may function as an early risk factor for dementia risk by modulating metabolic pathways and the neuro-immune axis.

## Results

These analyses were based on six independent cohort studies (**sTable 1**), two publicly available genomic datasets, other open-source references, and experimental observations in primary human macrophages (**Table 1**).

**Table 1.** Cohort and resource application summary The present analyses involved six independent cohorts and utilized multiple open-source resources for exploration, identification, and annotation.

| Data Set | PMID(s) | Type | Proteomics |  | Dementia |  |  |  |  |  |  | Imaging |  | Plasma<br>ADRD<br>Biomarkers | Genetics |  | Cell<br>culture | Annotation |
| --- | --- | --- | --- | --- | --- | --- | --- | --- | --- | --- | --- | --- | --- | --- | --- | --- | --- | --- |
|  |  |  | Plasma | CSF | Incident |  |  |  |  | Prevalent |  | MRI | PET |  | Instrum. | Sum.<br>Stats |  |  |
|  |  |  |  |  | ACD | AD | VaD | PDD | FTD | ACD | AD |  |  |  |  |  |  |  |
| ARIC | 2646917 | Cohort | ✓ |  | ✓ |  |  |  |  |  |  | ✓ |  | ✓ | ✓ |  |  |  |
| UKB | 34737426 | Cohort | ✓ |  | ✓ | ✓ | ✓ | ✓ | ✓ |  |  |  |  |  |  | ✓ |  |  |
| AGES-Reykjavik | 17351290 | Cohort | ✓ |  | ✓ | ✓ | ✓ |  |  |  |  |  |  |  |  |  |  |  |
| BLSA | 19126858 | Cohort | ✓ |  |  |  |  |  |  | ✓ |  | ✓ | ✓ | ✓ |  |  |  |  |
| NILS-LSA | 10835824 | Cohort | ✓ |  | ✓ |  |  |  |  |  |  |  |  |  |  |  |  |  |
| JHNC |  | Cohort | ✓ | ✓ |  |  |  |  |  |  |  |  |  |  |  |  |  |  |
| BioFinder | 39187705 | OSR |  | ✓ |  | ✓* |  |  |  |  | ✓ |  |  |  |  |  |  |  |
| deCODE genetics | 34857953 | OSR |  |  |  |  |  |  |  |  |  |  |  |  | ✓ |  |  |  |
| EADB | 35379992<br>23562540 | OSR |  |  |  |  |  |  |  |  |  |  |  |  |  | ✓ |  |  |
| FinGenn | 36653562 | OSR |  |  |  |  |  |  |  |  |  |  |  |  |  | ✓ |  |  |
| Apheresis Donors |  | BS |  |  |  |  |  |  |  |  |  |  |  |  |  |  | ✓ |  |
| Human Protein Atlas<sup>a</sup> | 32139519 | OSR |  |  |  |  |  |  |  |  |  |  |  |  |  |  |  | ✓ |
| ONTIME<sup>b</sup> | 34239129 | OSR |  |  |  |  |  |  |  |  |  |  |  |  |  |  |  | ✓ |
| Open Targets Genetics<sup>c</sup> | 33045747 | OSR |  |  |  |  |  |  |  |  |  |  |  |  |  |  |  | ✓ |
| Erichr<sup>d</sup> | 27141961 | OSR |  |  |  |  |  |  |  |  |  |  |  |  |  |  |  | ✓ |

Abbreviations: ACD, all-cause dementia; AD, Alzheimer's disease; ADRD, Alzheimer's disease and related dementias; AGES-Reykjavik, Age, Gene/Environment Susceptibility-Reykjavik Study; ARIC, Atherosclerosis Risk in Communities; BLSA, Baltimore Longitudinal Study of Aging; **BS, biological sample**; EADB, European Alzheimer & Dementia Biobank; FTD, Frontotemporal dementia; instrum., instrument; JHNC, Johns Hopkins Neurology Clinic; MRI, magnetic resonance imaging; NILS-LSA, National Institute for Longevity Sciences-Longitudinal Study of Aging; **OSR, open-source resource**; PDD, Parkinson's disease dementia; PET, positron emission tomography; PMID, PubMed Identifier; Sum. Stats, summary statistics; UKB, UK Biobank; VaD, vascular dementia
\*The BioFinder cohort was used to analyze protein levels across a pseudotime course of AD pathology
<sup>a</sup>Used to quantify protein cell and tissue expression; proteinatlas.org
Resources for single nucleotide polymorphism (SNP) summary statistics; <sup>b</sup>ontime.wustl.edu, <sup>c</sup>genetics.opentargets.org
<sup>d</sup>Public web tool used for gene set enrichment analysis; maayanlab.cloud/Enrichr

### Validation of plasma GDF15 measurements

We first confirmed the reliability of plasma GDF15 measurements on the SomaScan platform using previously reported results from independent cohorts. In a subsample of participants from the Atherosclerosis Risk in Communities (ARIC) study, SomaScan GDF15 measurements were strongly correlated with GDF15 measurements obtained from a proximity extension assay (Olink: *r*=0.79, *p*<0.0001, *N*=427) and two targeted immunoassays (Roche: *r*=0.92, *p*<0.0001, *N*=110; Luminex: *r*=0.83, *p*<0.0001, *N*=110)^15^ (**sFigure 1**, **sTable 2**). These findings are consistent with other large cohort studies that have confirmed the reliability of SomaScan GDF15 measurements using another immunoassay (R&D Systems)^16^ and proximity extension assays^17–19^ (**sTable 2**).

### GDF15 is broadly expressed outside the CNS and increases over the lifespan

To determine the origin of circulating GDF15 levels, we leveraged data from the Adult Genotype Tissue Expression Project (GTEx) Portal^20^ and Human Protein Atlas^21^. We found GDF15 is broadly expressed across many cell and tissue types, with the highest expression in the kidney, bladder, and choroid plexus (**sFigure 2; sTable 3**). However, there was limited GDF15 expression within CNS tissue or cells.

To validate GDF15’s previously reported association with older age^5,22^, we utilized participant data from the Baltimore Longitudinal Study of Aging (BLSA; *N*=1,313), who ranged in age from 22 to 93 years old.

Here, plasma GDF15 showed a strong, positive cross-sectional association with chronological age (*R*^2^=0.507, *p*<0.001; **Figure 2a**). Thus, although GDF15 increases are largely tied to age, approximately half of the variation in circulating GDF15 abundance is independent of age. GDF15 levels were more strongly correlated with age among males (*R*^2^=0.56) than among females (*R*^2^=0.46; age*sex interaction *p*=0.005), whereas other factors, such as obesity, did not affect GDF15’s association with age (**Extended Data Figure 1**). Plasma GDF15 showed a similar association with an epigenetic measure of biological age (DNAmPhenoAge; N=186, *R*^2^=0.40, *p*<0.001; **Figure 2b**). However, in analyses restricted to older adults (age 65+), plasma GDF15 levels showed much weaker associations with age (ARIC, *R^2^*=0.086 [**sFigure 3**]; BLSA, *R^2^*=0.186 [**sFigure 4**]). In addition to demonstrating that GDF15 expression is not specific to the CNS, these results suggest that age-related increases in circulating GDF15 may be more prominent across the lifespan rather than in older adulthood.

**Figure 2.**
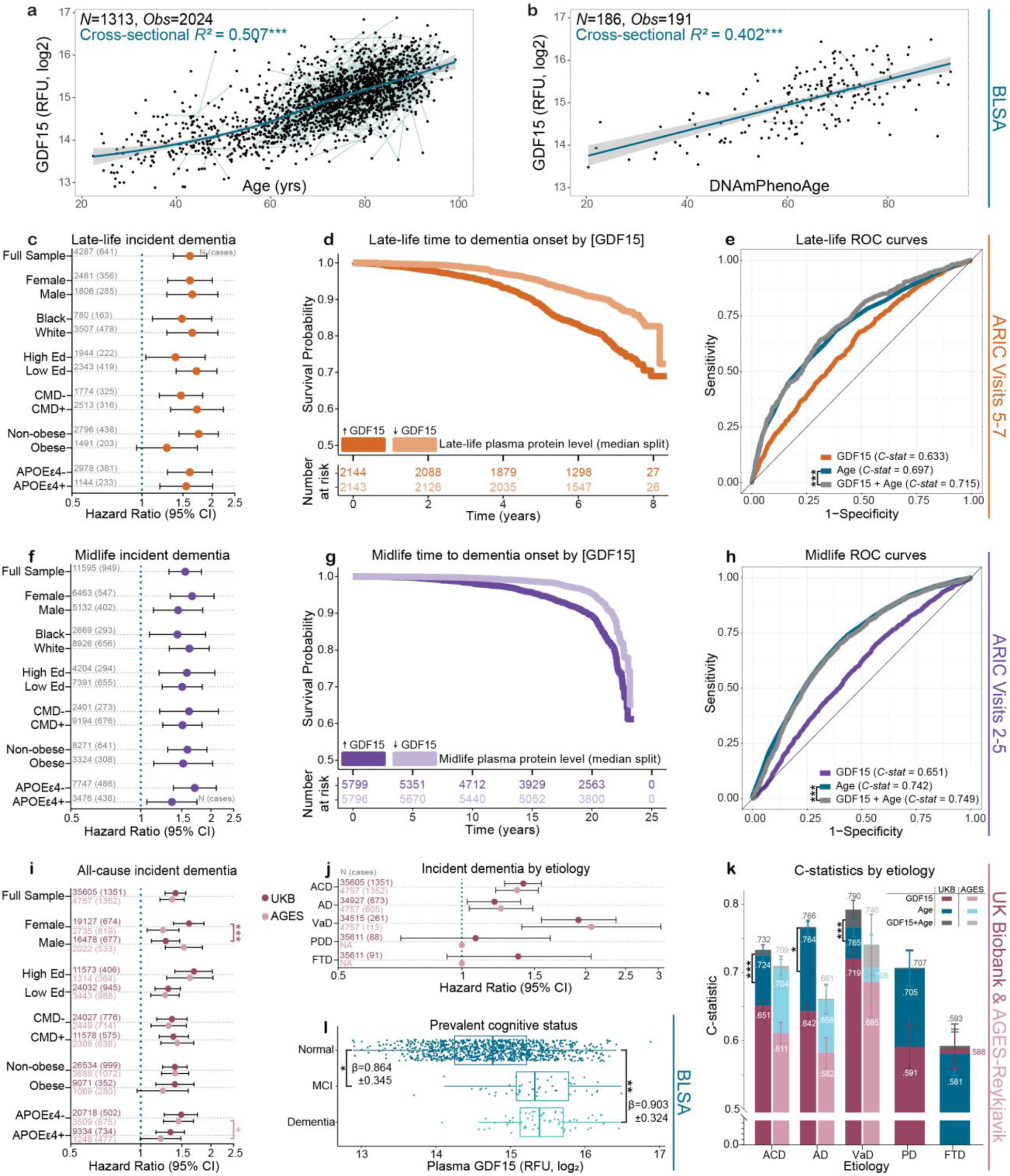
GDF15 abundance in plasma is associated with age and dementia risk across independent cohorts. **a.** Association of plasma GDF15 abundance and chronologic age in the BLSA. **b.** Association of plasma GDF15 and an estimation of phenotypic age based on DNA methylation (DNAmPhenoAge^100^). **c.** Associations of plasma GDF15 (measured in late-life) with 7-year all-cause dementia (ACD) risk in the full Atherosclerosis Risk in Communities (ARIC) study cohort and stratified by dementia risk factors, comorbidities, and demographic characteristics. **d.** Kaplan-Meier curves showing plasma GDF15 associations (high/low; median split) with ACD-free probabilities across late-life (7-year) in ARIC. **e.** ROC curves representing the classification of 7-year incident ACD status by late-life GDF15 levels alone, age alone, and GDF15 combined with age. **f.** Associations of plasma GDF15 (measured in midlife) with 20-year ACD risk in the full ARIC cohort and stratification by dementia risk factors, comorbidities, and demographic characteristics. ARIC results were obtained from Cox proportional hazards regression models adjusted for age, race-center, sex, education, APOEε4, eGFR, BMI, diabetes, hypertension, and smoking status. **g.** Kaplan-Meier curves showing plasma GDF15 associations (high/low; median split) with ACD free probabilities across midlife (20-year) in ARIC. **h.** ROC curves representing the classification of 20-year incident ACD status by midlife GDF15 levels alone, age alone, and GDF15 combined with age. **i.** Associations of plasma GDF15 with ACD risk in the UKB (14-year follow-up) and AGES-Reykjavik (17-year follow-up) cohorts and stratified by dementia risk factors, comorbidities, and demographic characteristics. **j.** Associations of plasma GDF15 with dementia etiologies in the full UKB and AGES-Reykjavik cohorts (PDD and FTD were not available in AGES). **k.** C-statistics representing plasma GDF15’s predictive performance for etiology specific dementia risk in the UKB and AGES-Reykjavik cohorts. Results were obtained from Cox proportional hazards regression models adjusted for age, sex, highest education, study site, BMI, eGFR, diabetes, and high cholesterol. **l.** Associations of plasma GDF15 with cognitive status in the Baltimore Longitudinal Study of Aging (BLSA). BLSA results were obtained from logistic regression models adjusted for age, sex, race, education, APOEε4, eGFR, and a comorbidity index. p-value: *<0.05, **<0.01 ***<0.001 *Abbreviations*: ACD, all-cause dementia; AD, Alzheimer’s disease; CMD, cardiometabolic disease; Ed, education; FTD, frontotemporal dementia; PDD, Parkinson’s disease dementia; ROC, Receiver Operating Characteristic; VaD, vascular dementia

### Higher plasma GDF15 is strongly associated with dementia risk in multiple large cohorts

To assess the relationships between plasma GDF15 and incident dementia, we analyzed proteomic data from three large cohort studies: ARIC (SomaScan v4.0), the UK Biobank (UKB; Olink Explore), and the AGES-Reykjavik study (SomaScan v4.1). To determine whether the association was modified by demographic characteristics, comorbidities, or established dementia risk factors, we conducted stratified analyses by sex, race, educational attainment (high/low; median split [years]), cardiometabolic disease (CMD; pos/neg), obesity (body mass index [BMI]≥30), and *APOE*ε4 carrier status (pos/neg).

In plasma collected from ARIC participants during late-life (*N*=4,287; mean age: 75.2±5; 58% female; 18% Black), higher GDF15 was associated with increased all-cause dementia (ACD) risk over a 7-year follow-up period (HR=1.61; 95% CI: 1.36, 1.90) after adjustment for demographic factors, physiological variables, and comorbid health conditions (**Figure 2c**). Unadjusted and minimally adjusted analyses yielded similar results (**sFigure 5a, sTable 4**). In stratified analyses, the association between GDF15 and ACD risk was consistent across subgroups, except among obese participants, where effects were attenuated (HR=1.28; 95% CI: 0.95, 1.73). Conversely, the strongest associations between GDF15 and dementia risk were observed in non-obese participants (HR=1.75; 95% CI: 1.44, 2.13). As illustrated in **Figure 2d**, ARIC participants with high plasma GDF15 levels in late-life (median split) showed a doubling of 7-year dementia risk compared to those with low GDF15 levels (18.7% vs 9.5% event rates, respectively; stratified results displayed in **Extended Data Figure 2**). With respect to 7-year dementia prediction, plasma GDF15 offered small, but statistically significant, improvements compared to age alone (C-statistics of 0.697 vs 0.715; **Figure 2e**, **sTable 4**).

Given the robust associations of plasma GDF15 with late-life ACD risk in older adults, we then examined the association of GDF15 with long-term (20-year) dementia risk using plasma collected from ARIC participants during midlife (*N*=11,595; mean age: 57.1±6; 56% female, 23% Black). Consistent with our prior results from a later ARIC visit^2^, each fold increase in midlife GDF15 was associated with a 55% increased risk of ACD risk over a 20 year follow-up (HR=1.55; 95% CI: 1.32, 1.82; **Figure 2f**, **sFigure 5b, sTable 4**). These results were maintained when analyses were restricted to participants who had GDF15 measured in early midlife (age<55; *N*=4,365; HR=1.57; 95% CI: 1.05, 2.34). Furthermore, midlife plasma GDF15 abundance was associated with ACD risk across all subgroups, with the strongest associations observed among *APOEε4*-negative participants (HR=1.71; 95% CI: 1.38, 2.11). ARIC participants with high plasma GDF15 levels in midlife (median split) showed a doubling of 20-year dementia risk compared to those with low GDF15 levels (7.5% vs 3.9% event rates, respectively; **Figure 2g, Extended Data Figure 3**).

Compared to age alone, midlife GDF15 in combination with age again offered small, but statistically significant, improvements in predictive performance for 20-year ACD risk (C-statistics of 0.742 vs 0.749; **Figure 2h, sTable 4).**

As part of a cross-platform validation to identify plasma GDF15’s associations with etiology-specific dementia risk, we next analyzed Olink proteomic data from UKB participants (*N*=35,673; mean age: 60.7±5; 54% female; 1% non-White). Higher levels of plasma GDF15 were associated with increased ACD risk over a 14-year follow-up period (HR=1.41; 95% CI: 1.27, 1.56; **Figure 2i, sTable 5**). Although this relationship was maintained in all risk strata, it was significantly stronger in females, compared to males (interaction*-p*=4.7×10^-4^). With respect to dementia etiology, each log_2_ increase in plasma GDF15 abundance was associated with a 92% higher risk for vascular dementia (VaD; HR=1.92, 95% CI: 1.56, 2.37) and a 20% higher risk for AD (HR=1.20, 95% CI: 1.03, 1.39) over 14 years. No significant associations were detected with incident Parkinson’s disease dementia (PDD) or frontotemporal dementia (FTD; **Figure 2j**). Models that incorporated GDF15 and age significantly outperformed an age only model for prediction of 14-year dementia risk, especially for VaD (C-statistics of 0.765 vs 0.790; **Figure 2k**, **Extended Data Figure 4, sTable 5**).

The association between serum GDF15 and long-term ACD risk (up to 17-year follow-up) was additionally replicated in a group of 4,757 participants (mean age: 76±5; 57.5% female) from the AGES-Reykjavik study (HR=1.36; 95%CI: 1.21, 1.53; **Figure 2i**, **sTable 6**). In stratified analysis, this association was particularly strong in *APOE*ε4-negative compared to *APOE*ε4-positive participants (interaction*-P=*0.043; **Figure 2i**, **sTable 6**). Similarly, GDF15’s relationship with increased dementia risk was consistently stronger among *APOEε4*-negative participants in both the UKB and ARIC, although the moderating effect of *APOE* genotype did not reach statistical significance in these cohorts. Analyses of etiology-specific dementia risk in the AGES-Reykjavik study revealed that each log_2_ increase in serum GDF15 abundance was associated with a 106% higher risk for VaD (HR= 2.06, 95% CI: 1.40, 3.04) and a 24% higher risk for AD (HR=1.24, 95% CI: 1.04, 1.49) over up to 17 years (**Figure 2j**, **sTable 6**). Compared to age alone, GDF15 in combination with age was a better predictor of VaD (C-statistic of 0.706 vs 0.740), but not AD or ACD (**Figure 2k**, **sFigure 6, sTable 6**). Although robust and consistent associations of GDF15 with incident dementia were observed across European and American cohorts, this finding did not extend to a smaller Japanese cohort of 340 participants from the National Institute for Longevity Sciences-Longitudinal Study of Aging (NILS-LSA; N=340; mean age: 66.7±5.2, 54.1% female; HR: 1.19, 95% *CI*: 0.90-1.57, *p*=0.24). In cross-sectional analyses in the BLSA, plasma GDF15 was elevated among those with prevalent MCI (N=50) and dementia (N=63), compared to cognitively normal controls (N=1,019; mean age: 69.2±16, 53% female; **Figure 2l**). Notably, higher plasma GDF15 was previously associated with worse cognitive functioning across all domains, except memory, in the Generation Scotland cohort study^23^. Taken together, these findings demonstrate that individuals who have elevated GDF15 levels in plasma during midlife or late-life are at increased risk for developing dementia, specifically dementia of a presumed vascular etiology (VaD).

### Mendelian randomization post-selection inference (MR-SPI) causally implicates plasma GDF15 in dementia risk

To assess the potential causal role of GDF15 on ADRD, as well as associated endophenotypes, we employed the recently developed MR-SPI approach^24^, which selects protein quantitative trait loci (pQTLs) as genetic instruments and performs robust causal inference in a two-sample MR framework. For primary analysis, we used plasma GDF15 *cis*-pQTLs derived from the ARIC cohort^2^ (n=7,597) to proxy lifetime GDF15 exposure. Outcome data were sourced from five genome-wide association study (GWAS) summary statistics: ADRD (n=487,511; AD cases=39,106; AD proxy cases=46,828), CSF Aβ42, and CSF pTau-181 (n=13,116) from the European Alzheimer & Dementia Biobank (EADB) consortium^25,26^, and ACD (n=211,397; cases=7,395) and AD (n=111,471; cases=3,899) from the Finnish Biobank (FINBB)^27^. Our primary analysis revealed significant positive associations of genetically proxied plasma GDF15 abundance with ADRD in the EADB consortium (but not AD in FINBB), CSF Aβ42, and CSF pTau-181 (**Figure 3a**, **sTable 7**). Using a distinct set of GDF15 plasma *cis*-pQTLs identified in a large Icelandic study (deCODE)^28^, we replicated the significant positive association of GDF15 with ADRD in the EADB consortium. However, the associations of GDF15 with CSF Aβ42 and CSF pTau-181 were not replicated. Notably, several of the identified GDF15 plasma pQTLs were also CSF pQTLs, suggesting a genetic coregulation of GDF15 across biofluids (**Figure 3b**; **sTable 8**).

**Figure 3.**
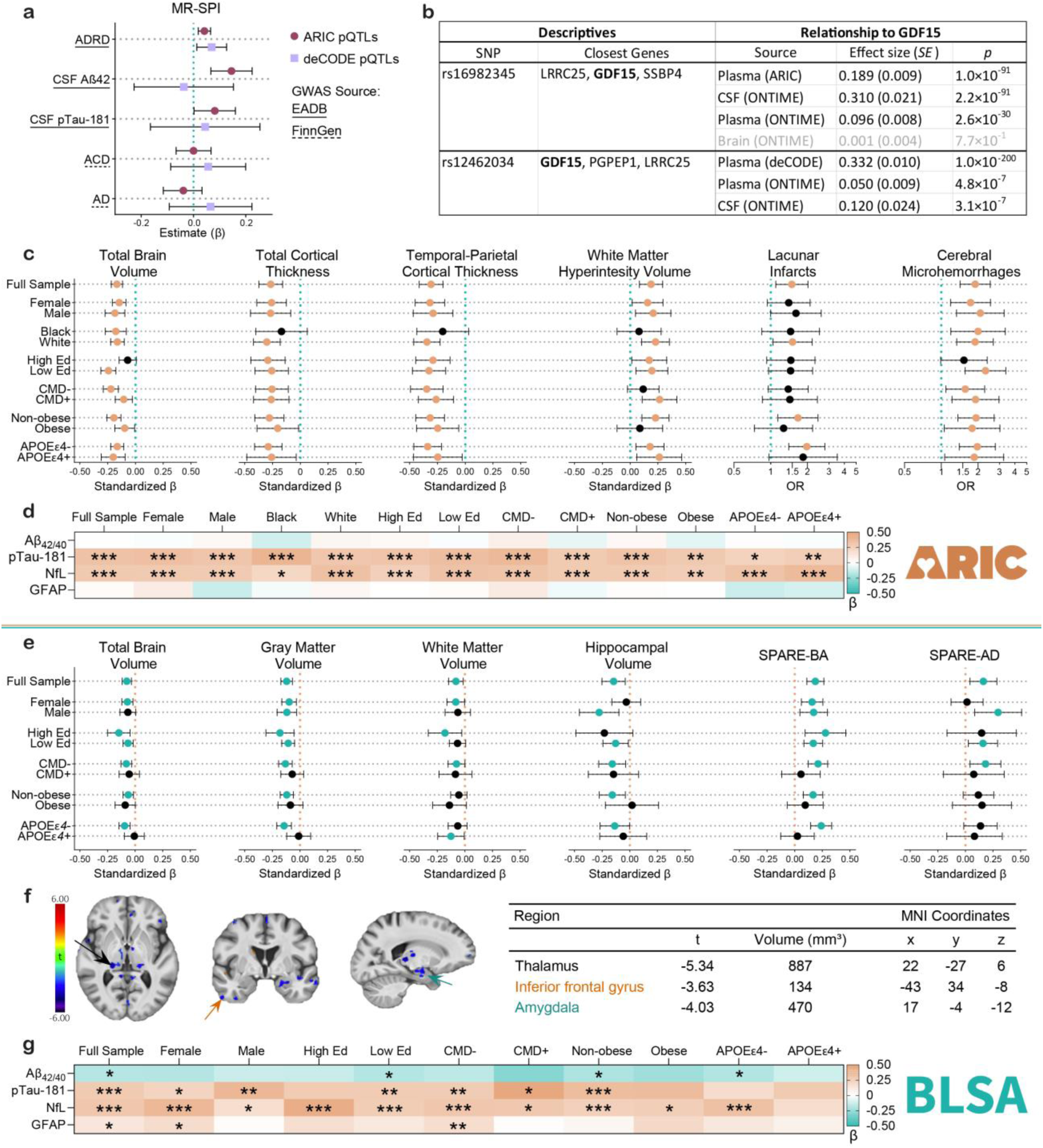
GDF15 abundance in plasma is associated with brain structure and plasma biomarkers across independent cohorts. **a.** Causal effect estimates were obtained using Mendelian randomization post-selection inference (MR-SPI) for clinically defined Alzheimer’s disease (AD) and related dementia (ADRD), all-cause dementia (ACD), cerebrospinal fluid measures of AD pathology and stroke. **b**. Top cis-pQTLs from the ARIC and deCODE studies and their association with plasma, CSF, and brain GDF15 abundance in other cohorts. **c.** Associations of plasma GDF15 with 3T MRI-derived structural brain imaging in the Atherosclerosis Risk in Communities (ARIC) study cohort and stratification by dementia risk factors, comorbidities, and demographic characteristics. **d.** Associations of plasma GDF15 with Alzheimer’s disease (AD) biomarkers in the ARIC cohort and stratified by dementia risk factors, comorbidities, and demographic characteristics. ARIC results were obtained from linear regression models adjusted for age, race-center, sex, education, APOEε4, eGFR, BMI, diabetes, hypertension, and smoking status; brain volume analyses also adjusted for intracranial volume. **e.** Associations of plasma GDF15 with 3T MRI-derived structural brain imaging in the Baltimore Longitudinal Study of Aging (BLSA) and stratification by dementia risk factors, comorbidities, and demographic characteristics. **f**. Associations of plasma GDF15 with voxel-wise brain volumes in the BLSA. An FDR-corrected threshold of p<0.05 was used to define significant clusters. **g.** Associations of plasma GDF15 with AD biomarkers in the BLSA cohort and stratification by dementia risk factors, comorbidities, and demographic characteristics. BLSA results were obtained from linear regression models adjusted for age, sex, race, education, APOEε4, eGFR, and a comorbidity index; brain volume analyses were also adjusted for intracranial volume. p: ***<0.001; **<0.01; *<0.05. *Abbreviations*: Aβ_42/40_, amyloid beta 42:40 ratio; CMD, cardiometabolic disease; EADB, European Alzheimer Disease Biobank; Ed, education; GFAP, glial fibrillary acidic protein; NfL, neurofilament light; pTau-181, phosphorylated tau 181.

### Plasma GDF15 is associated with neurodegeneration and cerebral small vessel disease

To characterize the neurobiological links between plasma GDF15 and dementia risk, we examined its associations with structural neuroimaging, fluid biomarkers, and Aβ positron emission tomography (PET). Among ARIC participants (*N*=1,345, mean age: 76.1±5.17, 40.8% female), each fold increase in plasma GDF15 was significantly associated with 0.17 standard deviation (SD) lower total brain volume, 0.27 SD lower total cortical thickness, 0.31 SD lower temporal-parietal cortical thickness, and 0.18 SD higher white matter hyperintensity (WMH) volume, as well as 40% and 90% higher odds of at least one lacunar infarct and cerebral microhemorrhage, respectively, in models adjusted for demographic factors, physiological variables, and comorbid health conditions (**Figure 3c**). We detected associations of similar magnitude for the hippocampus and a subcortical gray matter meta-region of interest (ROI; **sTable 9**).

In an independent sample of cognitively normal participants from the BLSA (*N*=994, mean age: 66.0±15, 55% female), GDF15 showed similar associations across ROIs (**Figure 3e**, **sTable 10**) and a diffused pattern of reduced gray matter volume (especially in the thalamus, amygdala, and inferior temporal gyrus) in voxel-based morphometry (**Figure 3f, sFigure 7, sTable 11**). Each fold-change increase in plasma GDF15 corresponded to a 0.16 SD lower volume in brain regions vulnerable to AD-related atrophy (SPARE-AD) and 0.19 SD lower volume in brain regions vulnerable to age-related atrophy (SPARE-BA), a pattern that we subsequently replicated in the UKB (N=2709, mean age: 63.7±8, 52% female; **Extended Data Figure 5, sTable 12**). An examination of five machine learning-derived predominant patterns of brain atrophy^29,30^ found that elevated plasma GDF15 was significantly associated with a pattern of diffuse cortical atrophy and perisylvian atrophy in BLSA, and a pattern of medial temporal atrophy and perisylvian atrophy in the UKB (**Extended Data Figure 5**). Notably, the pattern of perisylvian volume loss associated with GDF15 in both cohorts has been linked to MCI as well as dementia conversion, neuropsychiatric conditions, MS, PD, and an array of other age-related chronic diseases (e.g., reparatory, renal, metabolism, cardiovascular, etc).^31^ Together, these results show that, even before the onset of cognitive impairment, higher GDF15 levels in circulation accompany a pattern of enhanced neurodegeneration and cerebral small vessel disease.

With respect to fluid biomarkers^32^, higher plasma GDF15 was associated with plasma biomarker evidence of greater tau phosphorylation (pTau-181; standardized β=0.32) and neuronal injury (NfL; standardized β=0.29), but not soluble amyloid (Aβ_42/40_) or reactive astrogliosis (GFAP) in adjusted models (ARIC, N=1,411; **Figure 3d**, **sTable 13**). Similar associations of plasma GDF15 with pTau-181 (standardized β=0.28) and NfL (standardized β=0.30) were observed in cognitively normal BLSA participants (*N*=690 and 1017, respectively; **Figure 3g**, **sTable 14**), a finding which extended to CSF pTau-181 (β=0.433, SE=0.156, *p*=0.006) in a sample of 193 participants from the Johns Hopkins Neurology Clinic (JHNC; mean age: 55.3±21, 60.3% female). In contrast, plasma GDF15 was not associated with elevated cortical amyloid, as defined by PET imaging in ARIC (OR=0.90, 95% CI: 0.50, 1.63) or the BLSA (OR=1.22, 95% CI: 0.48, 3.07; **sTable 15**).

### Elevated plasma GDF15 is associated with neuro-immune activation in CSF

Using plasma and CSF proteomic data (SomaScan v4.1) from the JHNC, we found that GDF15 abundance in plasma was strongly correlated with GDF15 abundance in CSF (ρ=0.52, *p*=6.76×10^-5^) (**sFigure 8a)**.

Interestingly, this correspondence between plasma and CSF GDF15 was reduced in the context of dementia, but there was no change in correlation strength with respect to age (**sFigure 8b-c**). The high plasma-CSF GDF15 correlation is therefore unlikely to be driven by blood-brain barrier breakdown that can accompany increasing age and cognitive impairment. In this same cohort, plasma GDF15 correlated positively with select CSF measures of vascular dysfunction/inflammation (CDH5 [ρ=0.27], MMP12 [ρ=0.39], and VCAM1 [ρ=0.26]) and neuroimmune activation (TNFRSF1B [ρ=0.40], TREM1 [ρ=0.45], TREM2 [ρ=0.22], and LBP [ρ=0.31]; all uncorrected *p*<0.05; **Figure 4a**, **sTable 16**). CSF GDF15 showed a similar pattern of significant associations, albeit at a magnitude that was twice that of the plasma-to-CSF associations (e.g., VCAM1 [ρ=0.65], TNFRSF1B [ρ=0.80], TREM2 [ρ=0.67]; all uncorrected *p*<0.05). In a proteome-wide analysis of approximately 7000 proteins (SomaScan v4.1; uncorrected *p*<0.05), higher plasma GDF15 was associated with an altered abundance of plasma proteins involved in immune processes (e.g., IL-6/JAK/STAT3 signaling), ephrin receptor signaling, and extracellular matrix organization (**Figure 4b-c**, **sTable 17**), as well as an altered abundance of CSF proteins involved in complement activation and coagulation, IL-6/JAK/STAT3 signaling, *Staphylococcus aureus*, pertussis, and coronavirus infections, in addition to an attenuation in neuronal viability pathways (**Figure 4d-e**, **sTable 17**).

**Figure 4.**
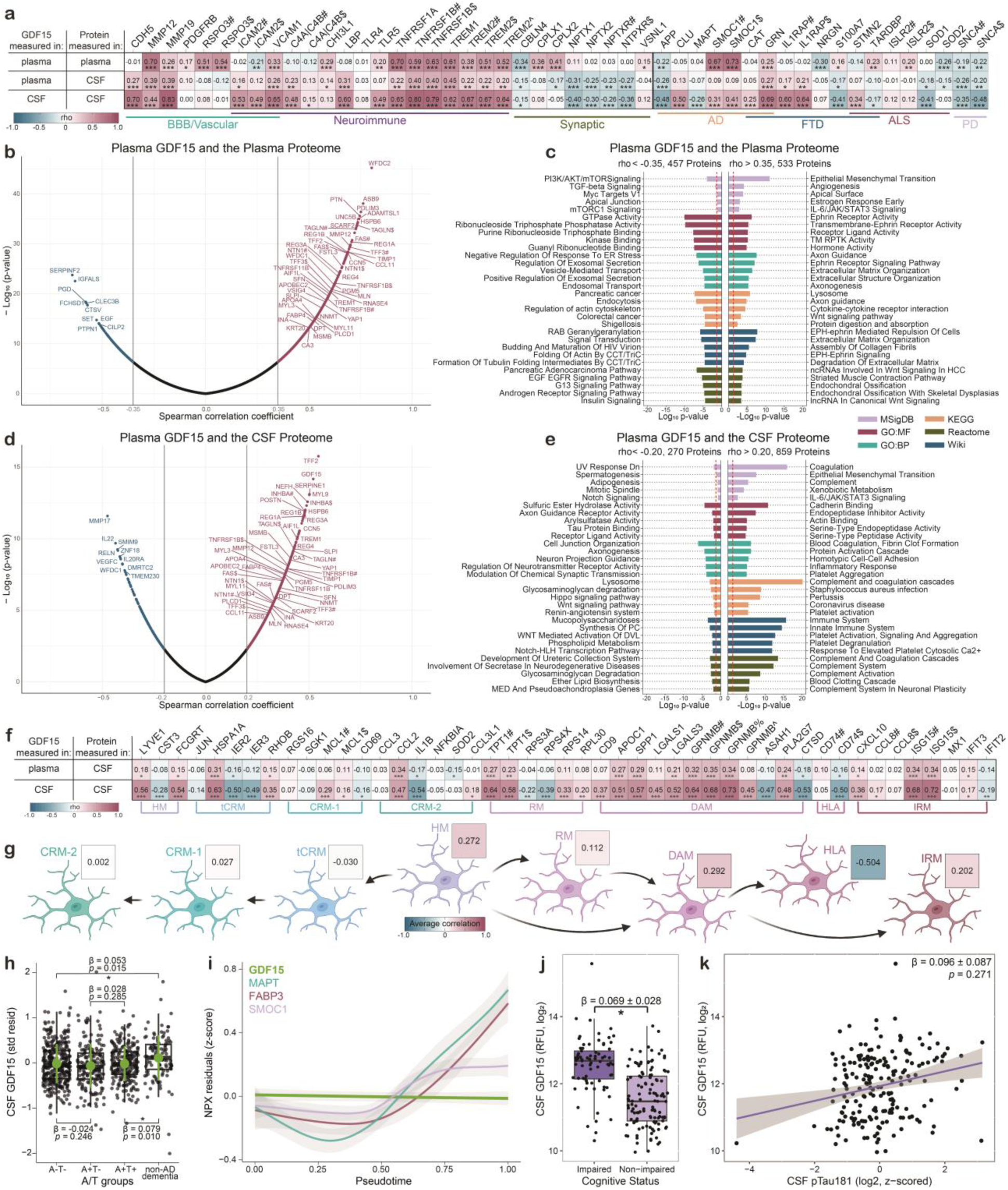
GDF15 relationships across and between biofluids (plasma and CSF). **a.** Heatmap depicting the Spearman correlations between GDF15 and select proteins previously associated with neurodegenerative disease and related neurobiological processes. **b.** Volcano plot of correlations between plasma GDF15 and the plasma proteome (SomaScan v4.1). **c.** Enrichment of plasma proteins which showed the strongest negative (left) and positive (right) correlations with plasma GDF15. **d.** Volcano plot of correlations between plasma GDF15 and the CSF proteome (SomaScan v4.1). **e.** Enrichment of CSF proteins which showed the strongest negative (left) and positive (right) correlations with plasma GDF15. **f.** Correlations of plasma and CSF GDF15 with CSF proteins whose cognate genes are differentially expressed in human microglial phenotypes. **g.** Diagram depicting the average Spearman correlations of CSF GDF15 with proteins whose cognate genes were differentially expressed in distinct microglial phenotypes, and the direction of microglial phenotypic stages associated with these correlations; if duplicate measures for the same proteins were present, the highest magnitude correlation was included in the average. **h.** Box plot depicting standardized residuals of CSF GDF15 levels among AD pathology subtypes (A+|-/T+|-) and non-AD dementia. **i.** Line plot depicting CSF protein level NPX residuals across pseudotime representing the course of AD. **j.** Box plot depicting levels of CSF GDF15 in cognitively impaired and cognitively unimpaired individuals in the JHNC Cohort (*N*=194); results were obtained from logistic regression adjusted for age and sex. **k.** Scatter plot depicting levels of CSF pTau181 (x) and CSF GDF15 (y) in the JHNC Cohort (*N*=193); results were obtained from linear regression adjusted for age and sex. *Abbreviations*: AD, Alzheimer’s disease; ALS, amyotrophic lateral sclerosis; BBB, blood-brain barrier; CRM, cytokine response microglia; CSF, cerebrospinal fluid; DAM, damage-associated microglia; FTD, frontotemporal dementia; HLA, antigen-presenting response; HM, homeostatic microglia; IRM, interferon response microglia; PD, Parkinson’s disease; RM, ribosomal microglia; tCRM, transitioning CRM

Given our results linking elevations in plasma GDF15 to immune pathways implicated in neuroinflammation (e.g., STAT3 and complement), we then asked whether GDF15’s CSF proteomic signature aligned with expression patterns obtained from single cell sequencing of different human microglia populations^33^. Specifically, we assessed the extent to which plasma and CSF GDF15 abundance correlated with CSF markers of homeostatic (HM), cytokine response (tCRM, CRM-1, and CRM-2), ribosomal (RM), disease-associated (DAM), human leukocyte antigen (HLA), and interferon response (IRM) microglia subtypes (**Figure 4f**, **sTable 17**). Plasma and CSF GDF15 showed strong associations (all uncorrected p<0.05) with several CSF proteins primarily expressed by CRM (HSPA1A, CCL2), DAM (APOC1, SPP1, GPNMB), and IRM (ISG15). Compared to plasma GDF15, intrathecal GDF15 was more strongly associated with these CSF markers of microglia subtypes, presumably due to the CNS specificity of genes that differentiate microglia populations. By averaging GDF15 correlations within specific microglial subpopulations, we found CSF GDF15 was most strongly related to CSF proteins implicated in the transition of microglia from a homeostatic to damage-associated and interferon-responsive state (HM-DAM-IRM pathway; **Figure 4g**).

### CSF GDF15 is elevated in the context of non-Alzheimer’s disease dementia and cerebral small vessel disease

To shed additional light on the role of CSF GDF15 in ADRD, we determined the extent to which CSF GDF15 is associated with biomarker-defined AD dementia, non-AD dementia, and cerebral small vessel disease using published data from the BioFINDER-2 cohort^24^ and new data from the JHNC. After adjusting for age, sex, and mean overall protein level, CSF GDF15 was not elevated among Aβ-positive/tau-negative (A+T-) participants compared to those without AD pathology (A-T-), or in Aβ-positive/tau-positive (A+T+) participants compared to Aβ-positive/tau-negative (A+T-) participants. However, CSF GDF15 was elevated among participants with non-AD dementia when compared to (i) participants with AD pathology (A+T+) and (ii) cognitively normal participants without AD pathology (A-T-; **Figure 4h**). In a pseudotime analysis that used CSF Aβ and tau to assign each BioFINDER participant along a continuum on the hypothesized AD course^24^, CSF GDF15 levels remained relatively constant, particularly in comparison to proteins that are known to increase with Aβ (A+/T-; SMOC1, MAPT) and tau (A+/T+; MAPT, FABP3; **Figure 4i**). Consistent with these results, CSF GDF15 was elevated in cognitively impaired (MCI or dementia), compared to cognitively unimpaired JHNC participants (β=0.069; SE=0.028; uncorrected *p*=0.02; **Figure 4j**), but was not associated with a CSF marker of phosphorylated tau (pTau-181; β=0.096, SE=0.087; uncorrected *p*=0.27; **Figure 4k**). CSF GDF15 has also been strongly associated with MRI indicators of cerebral small vessel disease, including higher WMH volume (β=0.09; uncorrected *p*=2.93×10^-7^) and lobar microbleeds (β=0.04; uncorrected *p*=0.02), in published results from the BioFINDER-2 cohort^34^. These associations of CSF GDF15 with cerebral small vessel disease but not AD pathology aligns with our results suggesting that GDF15 is more sensitive to VaD than to AD.

### Plasma GDF15 suppresses innate immune function and alters the human macrophage proteome

Because GDF15 abundance across biofluids maintained strong associations with neuroimmune processes despite its lack of CNS-specific expression patterns, we hypothesized that its mechanistic contributions to adverse neurological outcomes may be primarily mediated via peripheral immune mechanisms. To address this postulation and determine whether there is a causal link between circulating GDF15 protein levels and immune function, we leveraged several complementary approaches. Using GDF15 *cis*-pQTLs identified in ARIC and a two-sample MR framework^35,36^, we first examined relationships between genetically determined plasma GDF15 levels and 133 immune traits (**sTable 18**). This immuno-phenome-wide association study (IPheWAS) supported the causal relationship between elevated GDF15 and lower monocyte count (Z=-6.78; uncorrected *p*=1.14×10^-11^) and lower monocyte percentage (Z=-5.17; uncorrected *p*=2.37×10^-7^), after Bonferroni correction (**Figure 5a**). These findings provided proteogenetic support for GDF15’s inhibitory effect on innate immune processes.

**Figure 5.**
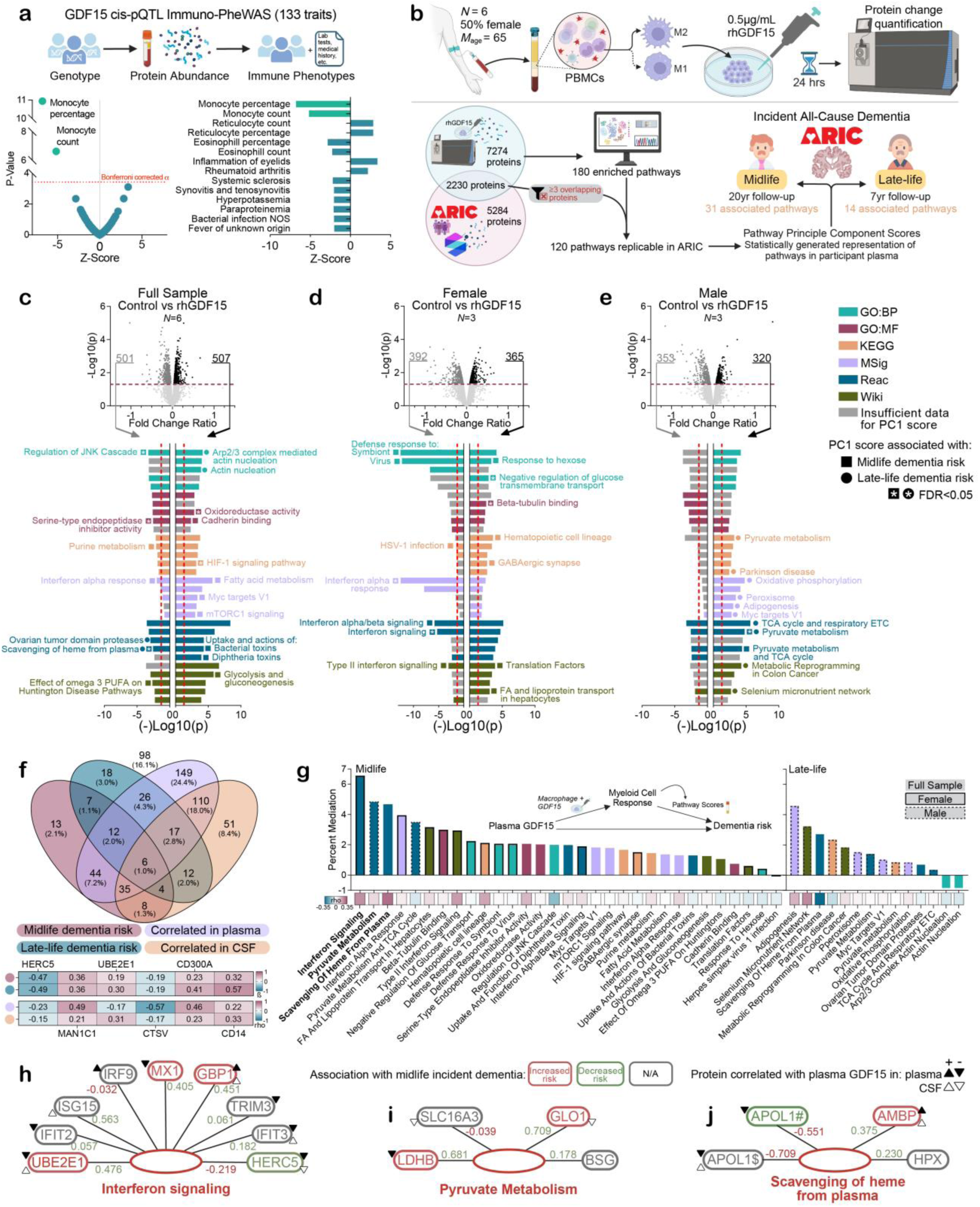
Probing GDF15 biology in primary human macrophages and translating results to clinical outcomes **a.** IPheWAS examining the relationship between genetically imputed GDF15 and 133 immune-related phenotypes. **b.** Top schematic depicts the experimental process of exposing PBMC-derive macrophages to rhGDF15. Bottom schematic depicts the analytic process of relating protein mass spectrometry results post-exposure to rhGDF15 to dementia risk in ARIC participants via first principal component score representation of pathways. **c.** Volcano plot of protein changes in PBMC-derived macrophages exposed to rhGDF15 (compared to controls); enrichment analyses using significantly altered proteins (p<0.05) are reflected in the corresponding bar charts. Analyses were repeated in **d.** females only and **e.** males only. Labeled pathways were associated with incident dementia in the ARIC study (midlife, 20yr follow-up; late-life, 7yr follow-up) using Cox proportional hazards regression models adjusted for age, race-center, sex, education, APOEε4, eGFR, BMI, diabetes, hypertension, and smoking status. **f.** Venn diagram depicting the number of proteins associated with rhGDF15 exposure in at least one of the prior analysis groups (a-c) and how they related to dementia risk (ARIC) and correlated with plasma GDF15 when measured in plasma and CSF (JHNC). The middle six proteins are thus significantly related to mid-and late-life dementia risk and correlated with plasma GDF15 when measured in both plasma and CSF; their association with each outcome is displayed in the heatmap below. **g.** Bar chart depicting the extent to which pathways component scores were estimated to mediate the association between plasma GDF15 an incident dementia in ARIC. **h, i, j.** Schematics depicting how proteins within the interferon signaling, pyruvate metabolism, and scavenging of heme from plasma pathways (respectively) related to midlife dementia risk and load onto first principal composite scores (ARIC). *Abbreviations*: ARIC, Arthrosclerosis Risk in Communities; IPheWAS, immuno-phenome-wide association study; PBMCs, peripheral blood mononuclear cells; PC1, first principal component; rhGDF15, recombinant human GDF15

Next, we determined how the manipulation of extracellular GDF15 protein levels (akin to altered GDF15 abundance in circulation) affects the proteomic profiles and biological processes of cultured human immune cells. Using macrophages differentiated from monocytes that were isolated from human PBMCs from six donors (mean age: 66.3±5; 50% female) in conjunction with mass spectrometry-based proteomics applied to cell lysates, we identified differentially abundant proteins following exposure to recombinant human GDF15 (rhGDF15; **Figure 5b**, **sTable 19**). Full sample and sex-stratified analyses were conducted because of the unique proteomic variance between male and female donors (**sFigure 9**). A total of 610 proteins were significantly (uncorrected p<0.05) influenced by treatment with rhGDF15 in the full sample, females only, and/or males only. Using these differentially regulated proteins for downstream enrichment analyses, we detected an elevation of proteins associated with pathogen exposure (e.g., bacterial toxins) and metabolic pathways (e.g., glycolysis and glucogenesis, mTORC1 signaling) and a reduction of proteins linked to interferon response and iron transport pathways **(Figure 5c**; **Extended Data Figure 6a, sTable 20**). In sex-stratified enrichment, we found further attenuation of immune pathways – particularly interferon signaling – in females (**Figure 5d**, **Extended Data Figure 6b, sTable 20**) and additional amplification of metabolic processes in males (**Figure 5e**, **Extended Data Figure 6c, sTable 20**). Of the proteins modulated by rhGDF15 exposure, 29 were associated with future dementia risk when measured during midlife and during late-life in ARIC, including six (HERC5, MAN1C1, UBE2E1, CTSV, CD300A, CD14) that showed significant correlations with plasma GDF15 when measured in both CSF and plasma in the JHNC (**Figure 5f**). Our analyses revealed that HERC5, an interferon-induced E3 protein ligase^37^, mediated 4% of the relationship between midlife GDF15 and 20-year dementia risk, whereas CD300A, an inhibitory immune receptor found on both myeloid and lymphoid cells^38^, mediated 6% of the relationship between late-life GDF15 and 7-year dementia risk (**sTable 21**). Given their consistent relationships with dementia risk and GDF15 levels across biofluids, these two proteins may play particularly important roles in mediating the biological mechanisms by which GDF15 influences dementia risk.

### Translating observed cell culture responses to dementia risk

To determine how plasma GDF15’s interactions with innate immune cells may influence dementia risk, we derived biologically informed composite (principal component) scores using proteins annotated in rhGDF15-macrophage-enriched pathways and determined the extent to which each pathway was associated with incident dementia over a 20-yr and 7-yr follow-up (ARIC; N=11,595 and N=4,287, respectively; **Figure 5b**, **sTable 22**). Out of 180 rhGDF15-macrophage-enriched pathways identified in our preceding experiments in culture (i.e., top five up and down regulated pathways identified with six public databases from the full sample, females only, males only), we were able to recapitulate 120 in ARIC using available plasma proteins measured by the SomaScan assay. Of these, 31 and 14 were associated with dementia risk when measured during midlife and late-life, respectively, in covariate-adjusted analyses (unadjusted p<0.05; **Figure 5c-e**). Dementia-associated, rhGDF15-macrophage-enriched pathways included a diverse set of immune (particularly interferon and viral/antiviral) and metabolic processes, the former being more prominent in female participants, and the latter in male participants.

Of particular interest were three rhGDF15-enriched pathways associated with higher 20-year dementia risk when assessed at midlife (FDR<0.05). These pathways—*interferon signaling*, *pyruvate metabolism*, and *scavenging of heme from plasma*—were identified as the strongest biological mediators of GDF15’s relationship with dementia risk (6.6%, 4.9%, and 4.7% mediation, respectively; **Figure 5g**, **sTable 23**). The interferon response pathway was downregulated in macrophages following exposure to rhGDF15 and positively associated with dementia risk when measured during midlife, as were three of the nine proteins annotated in this pathway (**Figure 5h**). These findings support GDF15’s immunosuppressive function, particularly with respect to antiviral signaling, and further suggest that plasma GDF15’s association with dementia risk is driven, at least in part, by its mitigation of myeloid cell interferon signaling (**Figure 6**). On the other hand, proteins involved in pyruvate metabolism were strongly upregulated in macrophages following exposure to rhGDF15 (particularly among male participants) and associated with elevated dementia risk when measured during midlife and late-life (**Figure 5i**). Proteins in this metabolic pathway – including glyoxalase I (GLO1) and lactate dehydrogenase B (LDHB) – modulate lactate/pyruvate levels and energy production, an excess of which is can promote macrophage infiltration into the perivascular regions of the CNS (e.g., choroid plexus), where they can then respond to pathogens and other damage-associated molecular patterns^39^ (**Figure 6a**). Proteins involved in the scavenging free heme (a cytotoxic byproduct of hemolysis) from plasma were downregulated in macrophages exposed to rhGDF15 and associated with dementia risk when measured during midlife and late-life (**Figure 5j**). By suppressing a network of heme scavenging proteins, such as APOL1 and AMBP, circulating GDF15 may promote extracellular increases in free heme, which can catalyze the formation of reactive molecules that cause neuronal damage, such as reactive oxygen species^40,41^ and oxidized low-density lipoproteins^42^ (**Figure 6a**).

**Figure 6.**
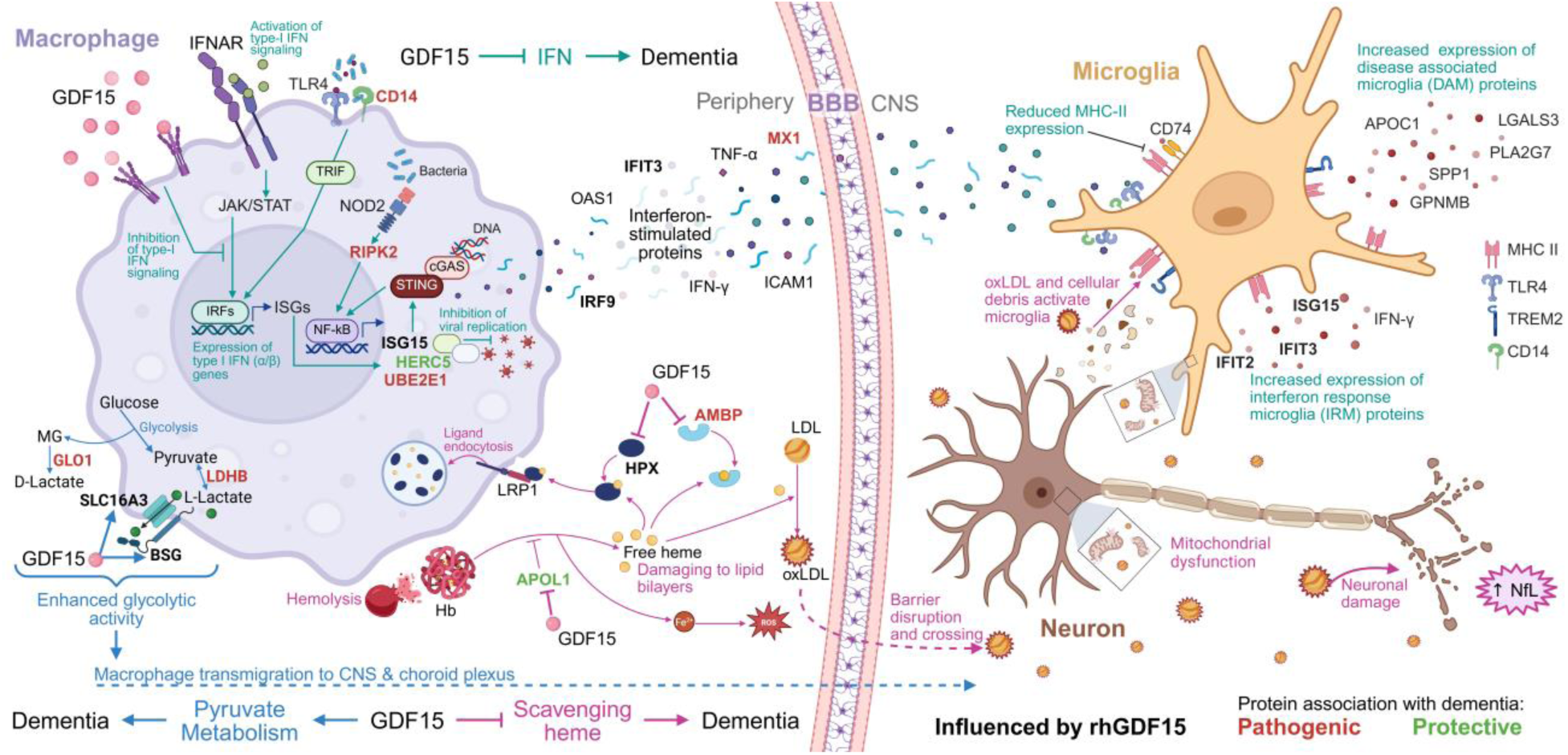
Proposed mechanisms linking GDF15 to dementia risk Proposed mechanisms of how pathways influenced by GDF15 exposure in macrophages (interferon response [teal], pyruvate metabolism [blue], scavenging of heme from plasma [pink]) may promote or protect against dementia risk.

## Discussion

Leveraging data from six independent cohorts, we demonstrated that elevated GDF15 in plasma, whether measured during midlife or late-life, is strongly associated with future dementia risk. GDF15 showed an association with VaD that was approximately two to five times greater than AD, highlighting its potential as a marker of vascular cognitive impairment. These findings were genetically supported, as results from MR analyses suggested a causal link between greater plasma GDF15 abundance and increased ADRD risk. Plasma GDF15 was also associated with a diffuse pattern of neurodegeneration and cerebral small vessel disease on MRI, elevated phosphorylated tau (as measured by pTau-181 in both plasma and CSF), and neuronal injury (as measured by NfL in plasma), even among cognitively unimpaired individuals. In contrast, plasma GDF15 was not associated with PET measures of cortical amyloid or plasma Aβ_42/40_, nor did it increase in CSF with progressive AD pathological staging. Individuals with elevated plasma GDF15 possessed plasma and CSF proteomic signatures indicative of immune and metabolic perturbations, a finding that was supported by our cell culture experiments, in which we treated primary human macrophages with rhGDF15. By relating biologic pathways modulated by rhGDF15 exposure to 20-and 7-year dementia risk, we identified specific physiological processes by which GDF15 may contribute to dementia pathogenesis, including an inhibition of the interferon/antiviral response, activation of pyruvate metabolism, and reduced scavenging of heme from plasma.

Although plasma GDF15’s association with dementia risk has been documented previously^2,12,22^, its utility as a biomarker and an understanding of its potential mechanistic role in brain aging and neurodegenerative disease have not been established. Here, we add to existing evidence by demonstrating that circulating GDF15 is elevated during midlife – before age 55 – in individuals at risk for dementia over the subsequent decades. A similar finding was previously reported by our group^2^ and confirmed here in an earlier (i.e., younger) wave of the ARIC cohort. GDF15 was associated with dementia risk in nearly all demographic and comorbidity strata, showing especially pronounced dementia associations among *APOE*ε4 non-carriers. This pattern aligns with plasma GDF15’s i) stronger association with VaD (compared to AD), ii) robust relationships with MRI indicators of cerebral small vessel disease, and iii) lack of association with PET cortical amyloid, plasma Aβ_42/40_, and progressive AD pathology. While we were able to replicate the putative causal link between plasma GDF15 and dementia (ADRD) risk using genetic (MR) analyses, our results, in total, suggest that this excess dementia risk is unlikely to be driven by GDF15’s effect on AD pathology (amyloid-β or tau), but rather through its effect on immune biology relevant to all forms of dementia, particularly VaD. Notably, the strong positive association between plasma GDF15 and pTau-181 (measured in plasma and in CSF) suggests GDF15 that (perhaps via the downstream immune changes) may play an early role in AD cascade via its link to soluble tau pathology.

By conducting an in-depth characterization of GDF15’s CSF proteomic signature, the current study sheds light on potential CNS mechanisms linking it to neurodegeneration. Our finding of a high correspondence between plasma and CSF GDF15 levels indicates that peripherally circulating GDF15 may migrate into the CNS. This may occur via the choroid plexus (i.e., where relatively high GDF15 expression is observed), or through alternative mechanisms, such as passive diffusion or active transport across the blood-brain barrier. Elevated GDF15 in plasma, in addition to being strongly associated with CSF GDF15, was associated with CSF evidence of neuro-immune activation. Specifically, proteins differentially expressed in the CSF of individuals with higher levels of plasma GDF15 were primarily enriched for complement and coagulation, IL6/JAK/STAT3 signaling, and infection related pathways. Further support for this link between plasma GDF15 and neuro-immune activation comes from our results which showed that GDF15 was associated with CSF levels of TREM2 and other proteins highly expressed by disease-associated (e.g., APOC1, SPP1, GPNMB) and interferon response (e.g., ISG15) microglial populations. CSF GDF15, which was elevated in the context of non-AD dementia, showed an even stronger association with DAM and IRM signature proteins compared to plasma GDF15, and a notable inverse association with C74, which is primarily expressed by antigen-presenting (HLA positive) microglial. These results suggest a cooccurrence of elevated GDF15 with specific expression profiles of microglia that are otherwise activated in response to damage-and pathogen-associated molecules, possibly through GFRAL (GDF15’s canonical CNS receptor)-independent mechanisms. Experimental studies are needed to definitively determine whether the link between plasma GDF15 and damage-responsive CSF immune signatures is the result of coordinated cross-talk of systemic and CNS immunological processes, despite its minimal expression in brain tissue and cell types.

While circulating GDF15 abundance correlates with molecular changes that are protective in some contexts (such as suppressing cancer cell proliferation^43,44^), results from our cohort analyses and two-sample MR support a pathogenic rather than protective role for GDF15 in dementia risk. Using primary human macrophages, we identified multiple biologically relevant protein networks and pathways that may mediate the dementia promoting effects of elevated plasma GDF15. Based on the size of these mediation effects, we prioritized investigation of interferon signaling, pyruvate metabolism, and heme scavenging pathways. Given our genetic and experimental evidence supports GDF15’s immunosuppressive effects, we suspect that GDF15 levels increase in response to interferon activity in an attempt to attenuate or resolve this innate antiviral response. In this scenario, either peripheral immune activation via interferon is protective, and the resulting GDF15-mediated immunosuppression increases dementia risk, or peripheral interferon signaling is pathogenic and GDF15 is a byproduct that influences dementia through alternative mechanisms. One such mechanism may be scavenging of heme from plasma, which we found to be downregulated by rhGDF15 and associated with increased dementia risk. This process appears to be driven, in part, by the decrease in APOL1 (i.e., a protein that helps prevent the creation of free heme following hemolysis) upon exposure to GDF15. Excess amounts of free heme are damaging to lipid bilayers and promotes the oxidization of low-density lipoproteins (LDLs).

Although intact LDLs do not normally cross the blood–brain barrier, its oxidized form may enter the brain, partly by contributing to barrier disruption^45^, giving rise to neuronal injury ^46^.

The present study has several strengths, including the use of multiple independent cohorts with up to twenty years of incident dementia follow-up, the implementation of high-throughput proteomic data in both blood and CSF, multimodal neuroimaging, and the use of cell culture experimentation to probe GDF15’s effect on immune cells. However, several limitations should be considered. First, because of the lack of CSF measurements available in our large cohorts with extensive follow-up, we were unable to determine if CSF levels of GDF15 might be a stronger predictor of dementia risk compared to intravenous levels. Second, although we used primary human macrophages to understand how GDF15 may affect the innate immune response, we did not perform these experiments with microglia; thus, we are unable to directly extrapolate how GDF15 may drive the neuro-immune axis. Third, not all proteins measured by mass spectrometry were also measured by the SomaScan assay (aptamer-based) used in our plasma proteomic measurements, and there can be discordance in protein measurements between these two techniques; therefore, the correspondence between the experiments in culture and the rhGDF15-macrophage-enriched pathways developed in the ARIC cohort may be limited. Despite these considerations, our findings provide insights into GDF15’s utility as a biomarker for cognitive impairment, and illuminate the biological mechanisms by which GDF15 may increase risk of dementia later in life.

## Methods

### Study design

We investigated how GDF15 protein levels in plasma relate to incident dementia and other endophenotypes of neurodegeneration via proteomic, neuroimaging, and genetic techniques. A multi-cohort study design (cohorts detailed below) was implemented to identify relationships of circulating GDF15 with incident dementia risk (ARIC, UKB, AGES-Reykjavik Study, NILS-LSA), neuroimaging outcomes (ARIC, BLSA), plasma biomarkers (ARIC, BLSA), and the plasma and CSF proteome (JHNC). We also examined whether demographic characteristics (e.g., self-reported sex) and other risk factors (e.g., presence of cardiometabolic disease) moderated these associations, applied genetic inference techniques, and employed cell culture conditions to determine the effects of manipulating GDF15 abundance. A detailed study design and flowchart is provided in **Figure 1**.

### The Atherosclerosis Risk in Communities (ARIC) study

#### Population

ARIC is an ongoing, community-based study that initially enrolled 15,792 mostly White and Black participants from four communities within the United States (Jackson, MS; northwestern suburbs of Minneapolis, MN; Forsyth County, NC; and Washington County, MD) between 1987 and 1989^47^. Until visit 4 (1996–1998), participants were evaluated every 3 years at in-person study visits. Visit 5 was then conducted in-person 15 years later (2011–2013), visit 6 took place about 5 years after (2016–2017), and visit 7 occurred about two years later (2018-2019). Data collection for subsequent ARIC study visits is ongoing. Non-White or non-Black participants were excluded, along with Black participants from Minneapolis and Washington Counties due to low sample sizes. Participants missing essential covariates, SomaScan protein measurements, or with a dementia diagnosis on or before the study’s baseline visit (visit 2 for midlife analyses, visit 5 for late-life analyses) were also excluded. Blood was drawn for proteomic analysis at visits 2 and 5. Midlife dementia risk was assessed between visits 2 and 5, and late-life dementia risk was assessed between visits 5 and 7. The neuroimaging and plasma biomarker data used in the present study were collected at visit 5. Institutional Review Boards (IRBs) approved the study protocols at each participating center: Johns Hopkins University, Baltimore, MD; University of Mississippi Medical Center, Jackson, MS; University of North Carolina at Chapel Hill, NC; and Wake Forest University, Winston-Salem, NC. All participants gave written informed consent at each study visit, and proxies provided consent for participants who were judged to lack capacity.

#### Plasma proteomics

Protein measurements were derived from blood samples collected at ARIC visits 2 and 5 using the SOMAmer-based array method (SomaScan v4.0 platform), as described previously^48^.

Plasma was collected using standardized protocols and frozen at −80°C until analysis. Using blind duplicates, the intra-assay coefficient of variation (CV) for GDF15 was 8.9% for visit 2 (*N*=520) and 10.7% for visit 5 (*N*=204). Due to skewed distributions among some protein measurements, all protein values were log_2_ transformed and those beyond 5 SDs were winsorized. Plasma collected at visit 5 was also used to validate SomaScan GDF15 quantification using an enzyme linked immunosorbent assay, as described previously^15^.

#### Targeted plasma biomarkers

Aβ_40_, Aβ_42_, GFAP, and NfL concentrations were measured using the Single Molecule Array (Simoa) Neurology 4-Plex E (N4PE). pTau-181 was measured using Simoa HD-X instrument (Quanterix) assays. Using 90 blind duplicates, CVs were 8.5, 7.3, 11.3, 9.8 and 9.7% for Aβ_40_, Aβ_42_, GFAP, NfL, and pTau-181, respectively. Values for GFAP, NfL, and pTau-181 were log_2_ transformed to correct for skewness. Aβ_42/40_ ratio and standardized values were used in analyses, and those beyond 5 SDs were winsorized. Biomarkers used in the present study were acquired at visit 5.

#### Dementia assessment

*Late-life analysis (visit 5 to 7).* At visits 5, 6, and 7, participants underwent comprehensive cognitive and functional in-person assessments to quantify memory, language, processing speed, and executive function, as described previously^49^. Between these visits, a phone surveillance approach was implemented, where participants were administered the Six Item Screener (SIS), a brief cognitive assessment, annually^50^. If the participant received a low score on the SIS (or was not able to participate in the screening via phone), the Ascertain Dementia 8 (AD8^51^) was administered to the participant’s informant. For participants who attended visits 6 and 7, these measures were used to estimate the date of dementia onset. For participants who did not attend visit 6 and 7 (due to death or nonattendance), these measures along with hospital discharge codes and death certificate codes were used to define dementia diagnoses and date of dementia onset^52^. An algorithmic dementia diagnosis was initially defined when the following criteria were met: a score >5 on the Functional Activities Questionnaire (FAQ) or a Clinical Dementia Rating scale sum of boxes (CDR-SB) >3; two or more cognitive domain scores >1.5×SD below the normative mean; and previous evidence of decline on the cognitive battery of >0.055×SD per year, which approximates the rate of cognitive decline in cognitively healthy older adults^53,54^. All dementia diagnoses identified using the algorithm were confirmed by an expert committee of physicians and neuropsychologists based on diagnostic criteria from the National Institute on Aging (NIA), the Alzheimer’s Association^55^, and the Diagnostic and Statistical Manual of Mental Disorders (Fifth Edition)^49,56^.

*Mid-life analysis (visit 2 to visit 5).* Dementia surveillance methodology for ARIC visit 1 through 5 have been detailed previously^49,57^. At visit 2, the baseline visit for the present analysis, and visit 4, participants were administered three neurocognitive tests (delayed word recall, digit symbol substitution, and word fluency test). After visit 2, telephone follow-ups were conducted annually. For a subset of participants suspected of having dementia, modified versions of the CDR and the FAQ were administered to informants. For participants who attended visit 5, these measures were used to estimate the date of dementia onset. For participants who did not attend visit 5 (due to death or nonattendance), the TICSm, CDR, FAQ, hospital discharge codes, and death certificate codes were used to define dementia diagnosis and date of dementia onset.

#### Brain magnetic resonance imaging

Structural brain images were acquired using 3T MRI scanners (Siemens Verio [Maryland study center], Siemens Skyra [North Carolina study center], Siemens Trio [Minnesota study center], and Siemens Skyra [Mississippi study center]). T1-MPRAGE and T2-FLAIR scans were acquired for all participants. Details of MRI acquisition have been described previously^58^. T1-weighted scans were used to estimate brain volumes; region-of-interest (ROI) volumes were quantified using FreeSurfer software^59,60^. In this study, the following neuroimaging measures were examined: total brain volume, total and temporal-parietal meta-ROI cortical thickness, subcortical gray matter meta-ROI volume, hippocampal volume, white matter hyperintensity volume, lacunar infarcts, and total, lobar, and subcortical cerebral microhemorrhages. White matter hyperintensity volume and lacunar infarcts were quantified from T2 FLAIR images using a computer-aided segmentation program (FLAIR-histoseg)^61,62^; white matter hyperintensity volumes were log-transformed. Cerebral microhemorrhages were identified from T2* Gradient Echo (GRE) images.

#### Amyloid PET imaging

^18^F-florbetapir PET (20 min.) scans were acquired as described previously^63^. 50-70 minutes after an intravenous bolus injection of the radiotracer, scans were conducted (Siemens).

Standardized uptake value ratios (SUVRs) were calculated using cerebellar gray matter region as a reference. Mean cortical Aβ reflected average SUVRs of the orbitofrontal, prefrontal, and superior frontal cortices, lateral temporal, parietal, and occipital lobes, precuneus, and anterior and posterior cingulate regions of interest. Aβ PET status (+/-) was defined based on a median of 1.20 mean cortical SUVR.

#### Covariates

Participant education (less than high school/high school; general education diploma or vocational school/at least some college), race (Black/White), and sex (male/female) were recorded at enrollment. Because race and study center are highly confounded, a race-study center variable was used (White-Washington County/White-Forsyth County/Black-Forsyth County/White-Minneapolis/Black-Jackson). APOE (coded as 0 APOEε4 alleles/≥1 APOEε4 alleles/missing) was genotyped using the TaqMan assay (Applied Biosystems). All other covariates (estimated glomerular filtration rate (eGFR, CDK-EPI^64^), hypertension, diabetes, BMI, and smoking status) were assessed at visits concurrent with plasma proteomic measurements used in the current analyses.

#### Statistical analyses

We used Cox proportional hazards regression models to examine the associations of plasma GDF15 abundance with incident dementia risk adjusted for demographic variables (age, sex, race-study center, and education), APOEε4 status, eGFR, and cardiovascular risk factors (BMI, diabetes, hypertension, and smoking status). The proportional hazards assumption was tested by plotting Kaplin Meier survival curves and by computing and plotting Schoenfeld residuals (**sFigure 10**). Covariates that did not meet the proportional Hazards assumption were incorporated in sensitivity analyses as stratified variables or with a time interaction (covariate*time) to determine whether results differed from that derived in the primary analyses. Multiple linear regression models, adjusted for the covariates listed above, were used to relate GDF15 levels to plasma biomarkers. The same multiple linear regression models were implemented to relate GDF15 levels with cross-sectional neuroimaging measures, which additionally adjusted for intracranial volume. Analyses were conducted using R v4.4.2 and Stata, version 17.

### UK Biobank (UKB)

#### Population

We analyzed data from a subset of participants from the UKB study, a population-based cohort of over half a million individuals aged 37 to 73 years at study entry. Blood samples used for plasma proteomics were collected at participants’ first visit between 2006 and 2010. Participants with missing plasma proteomics or covariate data, those younger than 65 at the date of censoring, and those who developed dementia before study entry were excluded from analyses. The collection and use of UKB data are approved by the Northwest Multi-Center Research Ethics Committee (Research Ethics Committee reference 11/NW/0382). All participants provided informed consent to use their data, health records, and biological materials for research purposes. The present study was conducted under the UK Biobank application number 83534.

#### Plasma proteomics

Plasma proteins were measured using Olink Explore 3072, as described previously^65^. Data pre-processing followed standard UKB quality control (QC)procedures; normalized protein expression values below the lower limits of detection were preserved, and log_2_ transformed values were used in analyses. The median intra-assay CV for GDF15 was 6.5%.

#### Dementia diagnosis

Dementia diagnoses were ascertained from hospital inpatient records (Hospital Episode Statistics [HES]) for all participants, with a subset of participants (45%) additionally having primary care (General Practice [GP]) data. Diagnoses were determined from ICD-9 or ICD-10 codes in the whole cohort as well as Read v2 or Clinical Terms Version 3 for primary care data. Specific diagnostic codes for dementia were identified from the UK NHS National Institute for Health and Care Excellence Quality and Outcomes Framework Business Rules, version 37.0. Read v2 and CTV3 codes were converted to ICD-9 and ICD-10 codes using UK Biobank Resource 592 (clinical coding classification systems and maps). Further details on diagnostic codes and ascertainment are available in our recent publication^66^. We examined all-cause dementia (ACD) as well as Alzheimer’s disease (AD), vascular dementia (VaD), Parkinson’s disease dementia (PDD), and frontotemporal dementia (FTD). For those who developed dementia, time to event was the interval in years from study entry to the date of diagnosis. For those who were free of dementia, time to event was the interval in years from study entry to 31 October 2022 for those who did not have a death record, and date of death for those who were deceased.

#### Brain magnetic resonance imaging

T1-weighted MPRAGE scans were acquired on a 3-T Siemens Skyra as previously described^67^. Machine learning-derived neuroimaging measures of age-and AD-related brain atrophy were generated using the same methodology as described in the BLSA (see below).

#### Covariates

Information on sex (male/female), race (self-reported ethnic background as white/non-white), study site, and educational attainment (highest education attained) were collected via participant self-reports. APOE genotypes were inferred from variants rs7412 and rs429358 on the Affymetrix Axiom/BiLEVE microarray platform^68^. All other covariates (eGFR (CDK-EPI^64^)-creatinine, diabetes, BMI, and high cholesterol) were ascertained concurrently with plasma proteomic measurements used in the current analyses.

#### Statistical analyses

Cox proportional hazards regression models were used to estimate the association between GDF15 and risk of incident dementia diagnosis adjusted for age, sex, education, study site, BMI, eGFR, diabetes, and high cholesterol. Multiple linear regression models adjusting for age, sex, BMI, eGFR, hypertension, diabetes, smoking, APOEε4 status, and total household income were used to examine associations of GDF15 with brain atrophy measures. Race was not adjusted for in UKB analyses given that 94.6% of UKB participants are white^17^. R (4.3.1) was used for statistical analyses.

### AGES-Reykjavik Study

#### Population

A detailed description of the AGES-Reykjavik Study^69^ has been provided previously. Briefly, the AGES-Reykjavik Study is a prospective longitudinal cohort study of older Icelandic adults who were initially enrolled in the Reykjavik Study that was established in 1967. From 2002 to 2006, 5,764 participants previously enrolled in the Reykjavik study were re-examined for the first wave of the AGES-Reykjavik study. This baseline assessment was completed over three visits within a 4-to 6-week window. Participants underwent a comprehensive assessment which included a clinical examination, questionnaires, a battery of cognitive measures, an MRI scan, and a blood draw. The AGES-Reykjavik study was approved by the Icelandic Nation Bioethics Committee (VSN: 00–063), the Icelandic Data Protection Authority, Iceland, and the IRB for the NIA. Written informed consent was obtained from all participants.

#### Serum proteomics

Blood samples used for the current analyses were collected at baseline visit. Serum was prepared using a standardized protocol and stored at −80° C until analysis. The SomaScan v4.1 platform was used to measure proteins. SOMAmers evaluated in the current study passed quality control. Values were log_2_ transformed.

#### Dementia adjudication

Dementia classification at the AGES-Reykjavik baseline and 5-year follow-up visit was conducted using a three-step procedure, as described previously^69^. All participants were administered the Mini-Mental State Examination and the Digit Symbol Substitution Test. Participants who received a low score on either measure were administered a more comprehensive battery of cognitive measures. Participants who received a low score on the Trails A and B measures or the Rey Auditory Verbal Learning Test received an additional assessment, which included a neurologic examination and a proxy interview. Dementia diagnoses (including AD and VaD) at the AGES visits were adjudicated based on consensus during a conference that included a neurologist, geriatrician, neuropsychologist, and a neuroradiologist. DSM-IV criteria were used to diagnose dementia^70^. All AGES-Reykjavik participants were followed up for incident dementia through nursing home reports (Resident Assessment Instrument [RAI]) and death certificates. The follow-up time was up to 16.9 years (until December 2019). Diagnoses for all-cause dementia and AD from nursing home reports were based on intake examinations upon entry or standardized procedures carried out in all Icelandic nursing homes. Diagnosis of AD was established according to National Institute of Neurological and Communicative Diseases and Stroke–Alzheimer’s Disease and Related Disorders Association (NINCDS-ADRDA) criteria or according to International Classification of Diseases, 10th revision (ICD-10) code F00 criteria. Individuals with incident non-AD diagnosis from nursing home reports who also had any vascular dementia ICD-10 code (F01) in hospital records were considered as incident VAD cases in addition to those diagnosed at the AGES follow-up visit.

#### Covariates

Age, sex, education, and lifestyle variables were assessed using questionnaires at baseline. Education was categorized as primary, secondary, college or university degree. Smoking was categorized as current, former or never smoker. APOE genotyping was assessed using microplate array diagonal gel electrophoresis (MADGE). BMI and hypertension were assessed at baseline. BMI was calculated as weight (kg) divided by height squared (m2), and hypertension was defined as antihypertensive treatment or blood pressure > 140/90 mmHg. Type 2 diabetes was defined from self-reported diabetes, diabetes medication use, or fasting plasma glucose ≥7 mmol L−1. Serum creatinine was measured using a Roche Hitachi 912 instrument, and eGFR was derived with the four-variable modification of diet in renal disease (MDRD) study equation^71^.

#### Statistical Analyses

Cox proportional hazards regression models were used to examine the association between the serum GDF15 and incident dementia risk occurring between the baseline visit and December 2019. Models were adjusted for baseline age, sex, education, APOEε4, eGFR-creatinine, BMI, diabetes, hypertension, and smoking status. R (4.4.3) was used for statistical analyses.

### Baltimore Longitudinal Study on Aging (BLSA)

#### Population

The BLSA is an ongoing community-based longitudinal study of physiological and psychological aging^72^. Recruitment and enrollment for the BLSA have been previously described^73,74^. In these analyses, we included 3T MRI scans acquired in 2008-2010, at which point blood was drawn for proteomic and AD and related dementia plasma biomarker analyses. The BLSA protocol was approved by the Institutional Review Board (IRB) of the National Institute of Environmental Health Science, National Institutes of Health (NIH) (03AG0325), and all participants provided written informed consent prior to participation. Participants were eligible for inclusion if they had available proteomic and MRI or plasma biomarker data and were excluded based on missing data and baseline cognitive impairment. Participants with cognitive impairment were retained in analyses examining the associations of GDF15 with cognitive status.

#### Plasma proteomics

Proteins in plasma were measured using the SomaScan v4.1 platform, as described previously^75^. Using 102 blind duplicates, proteins with intra-assay CV >50% were excluded (*N*=20); the CV for GDF15 was 4.5%. Values were log_2_ transformed and those beyond 5 SDs were winsorized.

#### Other plasma biomarkers

Aβ_40_, Aβ_42_, GFAP, NfL and pTau-181 concentrations in plasma were measured using the Single Molecule Array (Simoa) Neurology 4-Plex E (N4PE) and pTau-181 (V2) assays. Samples were run in duplicate, and values were averaged. CVs were 1.5, 1.0, 4.4, 4.9, and 4.8% for Aβ_40_, Aβ_42_, pTau-181, GFAP, and NfL, respectively. Aβ_42/40_ ratio was used in analyses, and values for GFAP, NfL and pTau-181 were log_2_ transformed. Values were standardized and those beyond 5 SDs were winsorized.

#### Cognitive status adjudication

Adjudication of cognitive status in the BLSA has been described previously^74^. In brief, a participant’s serial clinical and neuropsychological data were reviewed at each consensus case conference if the participant had > 3 errors on the Blessed Information-Memory-Concentration test, or a total combined score > 0.5 on the Clinical Dementia Rating Scale. MCI was based on Petersen’s criteria, and dementia was based on the criteria outlined in the Diagnostic and Statistical Manual of Mental Disorders, third edition revised, and by the National Institute of Neurological and Communicative Disorders and Stroke-Alzheimer’s Disease and Related Disorders Association.

#### Brain magnetic resonance imaging

T1-MPRAGE scans were acquired on a 3-T Philips Achieva. The validated Multi-atlas Region Segmentation Utilizing Ensembles (MUSE) anatomic labeling method designed to achieve a consistent parcellation of brain anatomy in longitudinal MRI studies using T1-weighted sequences was applied^76^. Voxel-wise maps for different brain tissue types were calculated using the validated RAVENS methodology^77^. The regions of interest for this study were total brain, gray matter, hippocampal, and white matter volumes. A machine learning-based score known as Spatial Pattern of Abnormality for Recognition of Early AD (SPARE-AD) was calculated for all participants, which was trained and validated in the Alzheimer’s Disease Neuroimaging Initiative cohort and captures multi-variate difference in brain structure that accurately discriminate cognitively normal subjects from those with neurodegenerative disease, particularly AD^78–80^. In brief, it is computed by training a support vector machine classification model to distinguish cognitively normal from clinically diagnosed AD individuals using structural brain features and has been shown to discriminate between normal cognition and mild cognitive impairment (MCI) as well as predict conversion for such individuals to MCI and AD dementia, respectively^81–83^. A machine learning-based score known as Spatial Pattern of Abnormality for Recognition of Early Brain Ageing (SPARE-BA) was calculated similarly and estimates the biological age of a participant’s brain structure at the time of the MRI scan^84^; however only scans from healthy controls were used for training the SVM used to calculate SPARE-BA. Using volumes of MUSE-segmented brain regions as input features, a semi-supervised deep representation learning approach (Surreal-GAN) was trained to calculate R indices (R1, R2, R3, R4, R5). By distinguishing heterogenous brain volume differences between younger (<50 years old) and older (>50 years old) adults, this approach generates multiple, continuous, low-dimensional scores that reflect the co-expression level of respective brain atrophy dimensions, and accounts for simultaneous spatial and temporal disease heterogeneity within the same individual. These scores have been trained and validated in a diverse cohort across 11 studies (>49,000 participants), where they predicted age-related clinical traits and disease diagnoses^31^.

#### Amyloid PET imaging

^11^C-Pittsburgh compound-B (PiB) distribution volume ratios (DVR; 70 min.) were collected on a GE Advance or Siemens High Resolution Research Tomograph scanner immediately following an intravenous bolus injection of approximately 555 MBq of the radiotracer. DVRs were computed using cerebellar gray matter as a reference. Mean cortical Aβ reflected the average DVR values across the cingulate, frontal, parietal (including precuneus), lateral temporal, and lateral occipital regions, excluding the pre-and post-central gyri. Mean cortical DVR values were harmonized between the two scanners by leveraging longitudinal data available on both scanners for 79 participants. Aβ PET status (+/-) was defined based on a Gaussian mixture model threshold of 1.064 mean cortical DVR^85^.

#### DNA methylation

DNA methylation was assayed using DNA extracted from blood samples collected at visits with concurrent proteomic data. CpG methylation status were determined using the Illumina Infinium HumanMethylation450 BeadChip (Illumina Inc., San Diego, CA), per the manufacturer’s protocol. Background correction, normalization, and quality control of data were conducted using the minfi package. DNA methylation PhenoAge (DNAmPhenoAge) was calculated using blood-based clinical phenotypes and chronological age^86^. For further details, please see Kuo, et al. ^87^.

#### Covariates

Sex (male/female), race (white/non-white), and education level (years) were defined based on participant self-reports. APOEε4 carrier status (0 ε4 alleles/≥1 ε4 alleles/missing) was defined via PCR with restriction isotyping using the Type IIP enzyme HhaI or the Taqman method. eGFR-creatinine was defined at the time of blood sample collection using the CKD-EPI criteria^88^. Comorbid diseases which represent potential confounders were defined using a comorbidity index calculated as the sum (score range: 0-8; converted to a percentage to account for missing data) of eight conditions: obesity, hypertension, diabetes, cancer, ischemic heart disease, chronic heart failure, chronic kidney disease and chronic obstructive pulmonary disease^89^.

#### Statistical analyses

Logistic regression models adjusted for age, sex, race, education, eGFR, APOEε4 status, and a comorbidity index were used to examine associations of plasma GDF15 with prevalent cognitive status. Linear regression models adjusting for the aforementioned covariates were used to examine associations of GDF15 with brain volumes and plasma biomarkers. Brain volume analyses additionally adjusted for intracranial volume (defined at age 70). For VBM analyses, an FDR-corrected threshold of p<0.05 was used to define significant clusters (clustering threshold = 50 voxels). SPARE-AD,SPARE-BA, and R-index analyses were adjusted for the aforementioned covariates except for intracranial volume (ICV) because ICV-residualized values were used in initial calculations. Analyses were performed using R (4.2.3).

### Johns Hopkins Hospital Neurology Clinic Cohort

#### Plasma and CSF proteomics

CSF and plasma samples from the same participants were used for proteomic analyses using SomaScan v4.1 assay. Subjects referred to the Johns Hopkins Hospital Neurology Clinic Cohort consented to banking of residual CSF after clinical testing under an IRB approved protocol. Samples that did not pass SomaScan QC criteria were excluded (CSF *N*=0; plasma *N*=2).

Samples identified as outliers by SomaScan or by principal component analyses were excluded (CSF *N*=3; plasma *N*=1). CVs were calculated using QC pooled, matrix-matched sample replicates provided by SomaLogic to monitor overall assay performance. The paired plasma-CSF study design spanned 3 plates for each biological matrix (plasma or CSF) and included 3 matrix-matched QC sample replicates in each plate. CVs were calculated as CV=100*SD(RFU)/mean(RFU), where RFU is the SOMAmer intensity in Relative Fluorescence Units measured for all intra-and inter-plate QC samples. Aptamers with CVs > 50% were excluded (CSF *N*=15; plasma *N*=18). The median CV for GDF15 was 3.0% for CSF and 3.5% for plasma. Values were log_2_ transformed and those beyond 5 SDs were winsorized.

#### Statistical analyses

To relate levels of GDF15 to the full proteome as measured by SomaScan across biofluids, spearman correlation analyses were implemented. The R package ‘pspearman’ was used to calculate correlations and provide a two-tailed p-value estimation. Analyses were performed using R (4.3.0). We conducted over-representation analyses using the set of the proteins most strongly correlated with plasma GDF15 (absolute value rho=0.35 for plasma, 0.2 for CSF) using Enrichr, a public, web-based tool that performs gene set enrichment analysis on user input^90–92^. To make a directionality distinction, separate enrichment analyses were ran on up-and downregulated proteins. The top five most significant pathways from six public databases (Gene Ontology: Biological Process [GO:BP], Gene Ontology: Molecular Function [GO:MF], Kyoto Encyclopedia of Genes and Genomes [KEGG], MSig Database (MSigDB), Reactome, and Wiki Pathways) were recorded (i.e., 30 pathways per direction-separated analysis). A background gene set of all proteins measured by SomaScan v4.0 (concatenated and repeated terms removed) was used for all analyses.

### Exposing differentiated macrophages to rhGDF15

#### Cohort demographics

Apheresis packs were collected from normal donors (*N*=6) aged 60 – 72 years old. Normal donors are volunteers and have donated apheresis packs through the Cytapheresis of Volunteer Donors protocol (NIA protocol # 03-AG-N316). Donors provided informed consent for their donations and the protocol was approved by the Institutional Review Board of the National Institute of Environmental Health Science, NIH.

#### Isolation of peripheral blood mononuclear cells (PBMCs)

Peripheral blood mononuclear cells (PBMCs) were isolated from the cytapheresis packs by density gradient centrifugation using Ficoll-Paque Plus (GE Healthcare). After isolation, PBMCs were cryopreserved in freezing media consisting of 15% dimethyl sulfoxide (DMSO) (Sigma) in fetal bovine serum (FBS) (Life Technologies) at a concentration of 50 million PBMCs per mL. Cryopreserved PBMCs were stored in liquid nitrogen freezer.

#### Cell culture

Cryovials of human peripheral blood mononuclear cells (PBMCs) were taken out of the liquid nitrogen freezer and immersed at 37°C water bath with stirring until the media was partially thawed. Cell suspension was then transferred to 9 mL of cold RPMI 1640 media supplemented with 10% FBS and centrifuged at 456 xg for 10 minutes. Cells were resuspended in warm RPMI media at a concentration of 2 million PBMCs per mL and incubated at 37°C for 1 h in a humified atmosphere. After incubation cells were centrifuged, counted and resuspended at a concentration of 50 million PBMCs per 1 mL in Robosep buffer according to the manufacturer’s instructions. Monocytes were enriched from the PBMCs using immunomagnetic negative selection cell EasySep human monocytes isolation kit (StemCell Technologies) using the automated cell separator RoboSep according to the manufacturer’s instructions (StemCell Technologies). After enrichment, 1.5 x10^6^ monocytes were seeded in 2 mL of ImmunoCult™-SF Macrophage Medium (StemCell Technologies) and differentiated into macrophages using 50 ng/mL of human granulocyte-macrophage colony-stimulating factor (GM-CSF, R&D Systems) at 37°C humidifier. 7 days after seeding, cells were washed once with macrophage medium and then treated with 0.5 μg/mL of recombinant human GDF15 (rhGDF15, R&D Systems Inc.) for 24 hour; after the 24-h treatment, protein lysates were collected for downstream analysis.

#### Protein isolation

Cells were lysed using a denaturing buffer [50 mM HEPES, 2% sodium dodecyl sulfate (SDS)] supplemented with protease and phosphatase inhibitors (Thermo Fisher Scientific). After boiling and sonication, samples were stored at-80°C. Protein concentration for each cell extract was measured using commercially available 2-D quant kit (Cytiva); protein extraction efficiency and sample quality were confirmed using SyproRuby stained NuPAGE gels (Thermo Fisher Scientific). Lipids and detergents from protein extracts were removed using methanol/chloroform extraction protocol^93^, and purified proteins were resuspended in 30 μL of urea buffer [8 M Urea, 2 M Thiourea, 150 mM NaCl (Sigma)], reduced by 50 mM DTT at 36°C for 1 h, and alkylated with iodoacetamide (100 mM) at 36°C in the dark for 1 h. Urea buffer with samples was diluted 12 times using 50 mM ammonium bicarbonate buffer. Proteins were then digested for 18 h at 36°C using trypsin/LysC mixture (Promega) in 1:50 (w/w) enzyme to protein ratio, and the resulting peptides were desalted using C18 cartridge (Restek), dried and stored at −80°C until further processing.

#### Sample preparation

TMT16plex (Thermo Fisher) was used to perform semi-quantitative proteomics. Briefly, 100 μg of peptides from 12 cultured macrophage lysates were spiked with 200 fM of bacterial beta-galactosidase digest (SCIEX) and treated with TMT reagent as recommended by the manufacturer.

All labeled TMT samples were merged and fractionated using the standard basic pH fractionation method.

High-pH RPLC fractionation was performed using the Agilent 1260 bio-inert HPLC system [3.9 mm X 5 mm XBridge BEH Shield RP18 XP VanGuard cartridge and 2.1 mm X 250 mm XBridge Peptide BEH C18 column (Waters)]. Mobile-phase composition was as follows: 10 mM ammonium formate (pH 10) as buffer A and 10 mM ammonium formate and 90% ACN (pH 10) as buffer B^94^. TMT-labeled peptides were resolved using a linear organic gradient (5% to 50% B in 100 min). Initially, 80 fractions were collected every minute during each LC run. Later, five individual fractions were merged into 15 combined fractions (fractions 1, 16, 31, 46, 61= combined as fraction one, fractions 2, 17, 32, 47, 62 = combined fraction two and so on). Combined fractions were speed vacuum dried, desalted and stored at −80°C until final LC-MS/MS analysis.

#### Mass spectrometry

Purified peptide fractions were analyzed using the Vanquish Neo UHPLC System coupled to an Orbitrap Ascend Tribrid mass spectrometer (Thermo Scientific). Each fraction was loaded using 2% acetonitrile in water with 0.1% formic acid (flow rate 10 μl/min) for 5 min onto a trap column 9Acclaim PepMap 100 HPLC Column, 75 μm ID, 2 cm long, 3 μm C18, Thermo Scientific) and then separated using EASY-Spray PepMap Neo UHPLC Column (75 μm, 75 cm long, 2 μm C18, Thermo Scientific) at 250 nl/min flow rate. The linear organic gradient went from 2% to 35% B in 145 min with total LC method duration of 180 minutes. Mobile phases A and B consisted of 0.1% formic acid in water and 0.1% formic acid in acetonitrile, respectively. Tandem mass spectra were obtained using the Orbitrap Ascend Tribrid mass spectrometer with a heated capillary temperature of +320°C and spray voltage set to 2.3 kV. Full MS1 spectra were acquired from 375 to 1500 m/z at 240,000 resolution and maximum injection time set to auto mode with automatic gain control (AGC) set to 1×10^6^. Dd-MS2 spectra were acquired using 0.7 m/z isolation window with normal m/z range and fixed first mass of 110 m/z. MS/MS spectra were resolved to 45,000 with maximum injection time set to auto and standard AGC target set to 5×10^4^. Over 3 seconds of total cycle time most abundant ions were selected for fragmentation with 34% normalized high collision energy. A dynamic exclusion time was set to 60 s to discriminate against previously analyzed ions. The raw data generated from each sample fraction was converted to mascot generic format (mgf) using MSConvert software (ProteoWizard 3.0.20026) and then searched with Mascot 2.4.1 and X!Tandem CYCLONE (2010.12.01.1) using the SwissProt Human sequences from Uniprot (Version Year 2023, 20,300 sequences) database. Quantitative values were expressed as fold change relative to control samples.

#### Bioinformatic analysis

Analysis of the identified proteins was carried out following the standard workflow of the R package DEP, version 1.26.0^95^. Only proteins detected in at least two out of three replicates in at least one experimental condition were included for analysis. Data were normalized using the variance stabilization normalization (VSN) method with the function “normalize_vsn”, and missing values were imputed using random draws from a Gaussian distribution centered around a minimal value with the function “MinProb”. The differential protein expression analysis was performed based on linear models and empirical Bayes statistics using limma via function “test_diff” in the DEP package, with adjustment for variability associated with the donor.

### Deriving biologically informed composite scores

Enrichment of differentially expressed proteins in primary macrophages following treatment with rhGDF15 was conducted using Enrichr as described above. The top five most significant pathways from six public databases (GO:BP, GO:MF, KEGG, MSigDB, Reactome, and Wiki) were recorded (i.e., 30 pathways per direction-separated analysis). Out of 180 experimentally determined rhGDF15-enriched pathways (from three samples [full, female only, and male only]; 60 pathways per sample [30 up, 30 down]), 120 were able to be reproduced using ARIC plasma proteomic data (SomaScan). At least three proteins in a pathway had to have been measured by both mass spectrometry and SomaScan to be considered reproducible; UniProt IDs were used to determine which proteins were measured by each methodology. Using the set of ARIC SomaScan proteins assigned to each reproducible rhGDF15-enriched pathway, we ran a principal component (PC) analysis and used the first principal component of each pathway to create individualized pathway-specific PC scores using each participant’s measured protein levels. Once PC-scores were generated, they were then used as predictor variables in Cox proportional hazards regression models to examine their associations with incident dementia risk over 7yr and 20yr follow-up periods (adjusted for all ARIC covariates discussed previously).

### Two-sample Mendelian Randomization (MR)

To assess the potential causal effects of GDF15 on ADRD and associated endophenotypes, we employed the recently developed MR-SPI approach. MR-SPI is a two-sample MR method that first selects valid genetic instruments that satisfy core instrumental variable (IV) assumptions (IV relevance, IV independence, and exclusion restriction assumptions), and then performs post-selection inference for reliable causal findings. Specifically, under the *Leo Tolstoy’s Anna Karenina principle*, valid genetic instruments will form the largest group and provide similar ratio estimates. Applying the voting procedure, MR-SPI selects the largest group where genetic instruments mutually vote for each other to be valid. This voting procedure is particularly advantageous in pQTL studies with limited candidate genetic instruments, as it does not rely on any additional distributional assumptions on the genetic effects. Subsequently, MR-SPI uses a searching-and-sampling method to construct a robust confidence interval to mitigate the potential finite-sample IV selection error. By integrating selection of valid genetic instruments with construction of robust confidence interval, MR-SPI strengthens the reliability of causal inference in MR analysis. The MR-SPI package is available at https://github.com/MinhaoYaooo/MR-SPI. Here, we performed linkage disequilibrium (LD) clumping with a 10,000 kb window and R^2^=0.1 using PLINK software^96^. After removing highly correlated pQTLs for primary (ARIC) and secondary (deCODE) MR analyses, 10 and 37 *cis-*pQTLs were retained as candidate genetic instruments, respectively. By leveraging the LD matrix extracted from a reference panel for potentially weakly correlated pQTLs, joint association estimates were computed using GWAS summary statistics^97^. Incorporating these joint association estimates into MR-SPI allows for the inclusion of additional pQTLs, thereby enhancing statistical power to select valid IVs and detect causal effects.

### Immuno-phenome-wide association study (IPheWAS)

To assess the potential causal effects of GDF15 on immune traits, we performed an immuno-phenome-wide association study (IPheWAS). IPheWAS was conducted using the FUSION pipeline based on the previously defined cis-pQTL predictions models from individuals of European ancestry in the ARIC study^35^ against immuno-related GWAS summary statistics datasets from the UK Biobank^98,99^ covering a total of 133 immune traits. To control for multiple testing, a Bonferroni correction was applied based on the number of aptamers modelled across the number of UK Biobank traits (p<3.76× 10^-4^ = 0.05 / 133 immune traits).

## Data availability

Data generated in the current study are available in this article (and its supplementary figures or supplementary tables), upon reasonable request, or in an online public repository. ARIC proteomic data is available through the National Heart, Lung, and Blood Institute (NHLBI) Biologic Specimen and Data Repository Information Coordinating Center (https://biolincc.nhlbi.nih.gov/studies/aric/). Additional requests for clinical or proteomic data may be submitted to the ARIC Steering Committee and will be reviewed to ensure that data can be shared without compromising participant confidentiality or breaching intellectual property restrictions. Participant-level demographic, clinical and proteomic data may be partially restricted based on prior participant consent, and data sharing restrictions may also be applied to ensure consistency with confidentiality or privacy laws and considerations (https://sites.cscc.unc.edu/aric/). Anonymized BLSA data not published within this article may be shared upon request from qualified investigators. Researchers who wish to use BLSA data are encouraged to develop a pre-analysis plan that can be submitted for approval (https://blsa.nia.nih.gov/how-apply). Data, protocols, and other metadata of the UKB are available to the scientific community upon request in accordance with the UKB data sharing policy (https://www.ukbiobank.ac.uk/enable-your-research/apply-for-access).

## Code availability

No original code was developed for this study, and no custom code or mathematical algorithm was central to its conclusions.

## Acknowledgements

The authors thank the staff and participants of the ARIC, UKB, AGES-Reykjavik, BLSA, NLS-LSA, and JHNC studies for their important contributions.

## Contributions

**Conceptualized study design**: Cassandra O. Blew, Michael R. Duggan, Dimitrios Tsitsipatis, Allison B. Herman, Keenan A. Walker. **Prepared data and performed statistical analyses:** Cassandra O. Blew, Michael R. Duggan, Gabriela T. Gomez, Zulema Rodriguez Hernandez, Luke C. Pilling, Jingsha Chen, Heather E. Dark, Yifei Lu, Cassandra M. Joynes, Eva Jacobsen, Minhao Yao, Zulema Rodriguez Hernandez. **Provided or prepared data for their respective studies:** Shannon M. Drouin, Abhay Moghekar, Julián Candia, Murat Bilgel, Aditi Gupta, Krystyna Mazan-Mamczarz, Myriam Gorospe, Alexey Lyashkov, Yevgeniya Lukyanenko, Lori L. Jennings, Valborg Gudmundsdottir, Vilmundur Gudnason, Lenore J. Launer, Naoto Kaneko, Shintaro Kato, Makio Furuichi, Masaki Shibayama, Masahisa Katsuno, Keita Hiraga, Yukiko Nishita, Rei Otsuka, James R. Pike, Mary R. Rooney, Guray Erus, Mary Kaileh, Iwao Waga, Michael Griswold, Christie Ballantyne, Yuhan Cui. **Obtained funding for elements of study**: Lori L. Jennings, Valborg Gudmundsdottir, Vilmundur Gudnason, Christie Ballantyne, Rebecca F. Gottesman, Priya Palta, Myriam Gorospe, Christos Davatzikos, Luigi Ferrucci. **Provided key statistical genetics guidance**: Minhao Yao, Zhonghua Liu, Zulema Rodriguez Hernandez, Pascal Schlosser, Keenan A. Walker. **Drafted initial manuscript**: Cassandra O. Blew, Michael R. Duggan, Keenan A. Walker. **Contributed to manuscript preparation or revisions:** Dimitrios Tsitsipatis, Gabriela T. Gomez, Zulema Rodriguez Hernandez, Luke C. Pilling, Heather E. Dark, Shannon M. Drouin, Cassandra M. Joynes, Minhao Yao, Murat Bilgel, Abhay Moghekar, Qu Tian, Julián Candia, Myriam Gorospe, Mika Kivimaki, Philipp Frank, Valborg Gudmundsdottir, Vilmundur Gudnason, Lenore J. Launer, James R. Pike, Mary R. Rooney, Pascal Schlosser, Yuhan Cui, Guray Erus, Christos Davatzikos, Rebecca F. Gottesman, Iwao Waga, Priya Palta, Christie Ballantyne, Michael Griswold, Zhonghua Liu, Luigi Ferrucci, Allison B. Herman

---

**Extended Data Figure 1.**
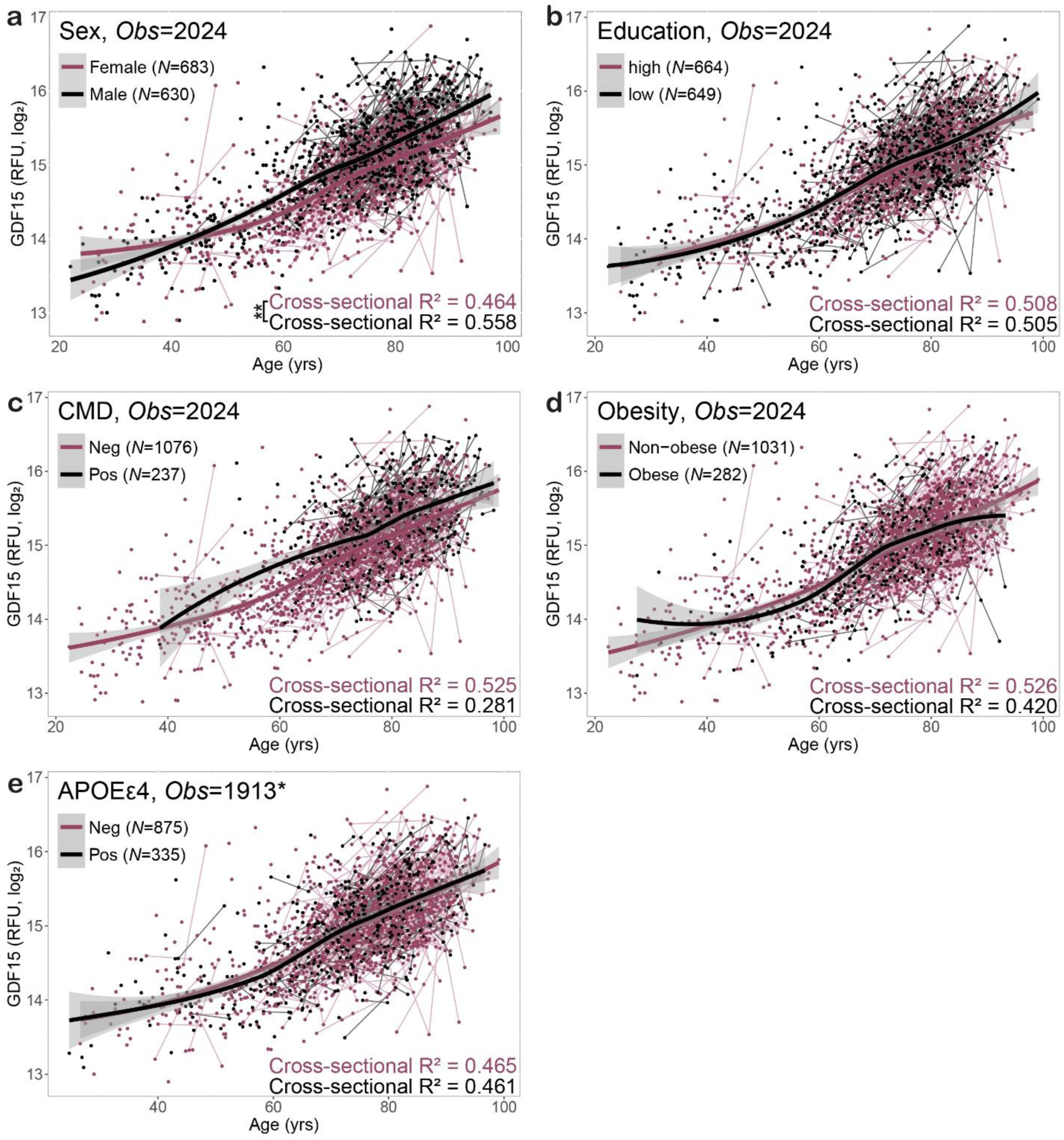
Plasma GDF15 associations with age among all participants (ages 22-99) in the Baltimore Longitudinal Study of Aging (BLSA). Associations of plasma GDF15 with age stratified by **a.** sex, **b.** educational attainment, **c.** cardiometabolic disease, **d.** obesity, and **e.** AD genetic risk (*APOEε*4 status; *Includes fewer observations due to unknown genotype). Results were obtained from unadjusted linear regression models which used the earliest available blood sample per participant; lines shown are loess curves. *Abbreviations*: AD, Alzheimer’s disease; CMD, cardiometabolic disease; Obs, observations

**Extended Data Figure 2.**
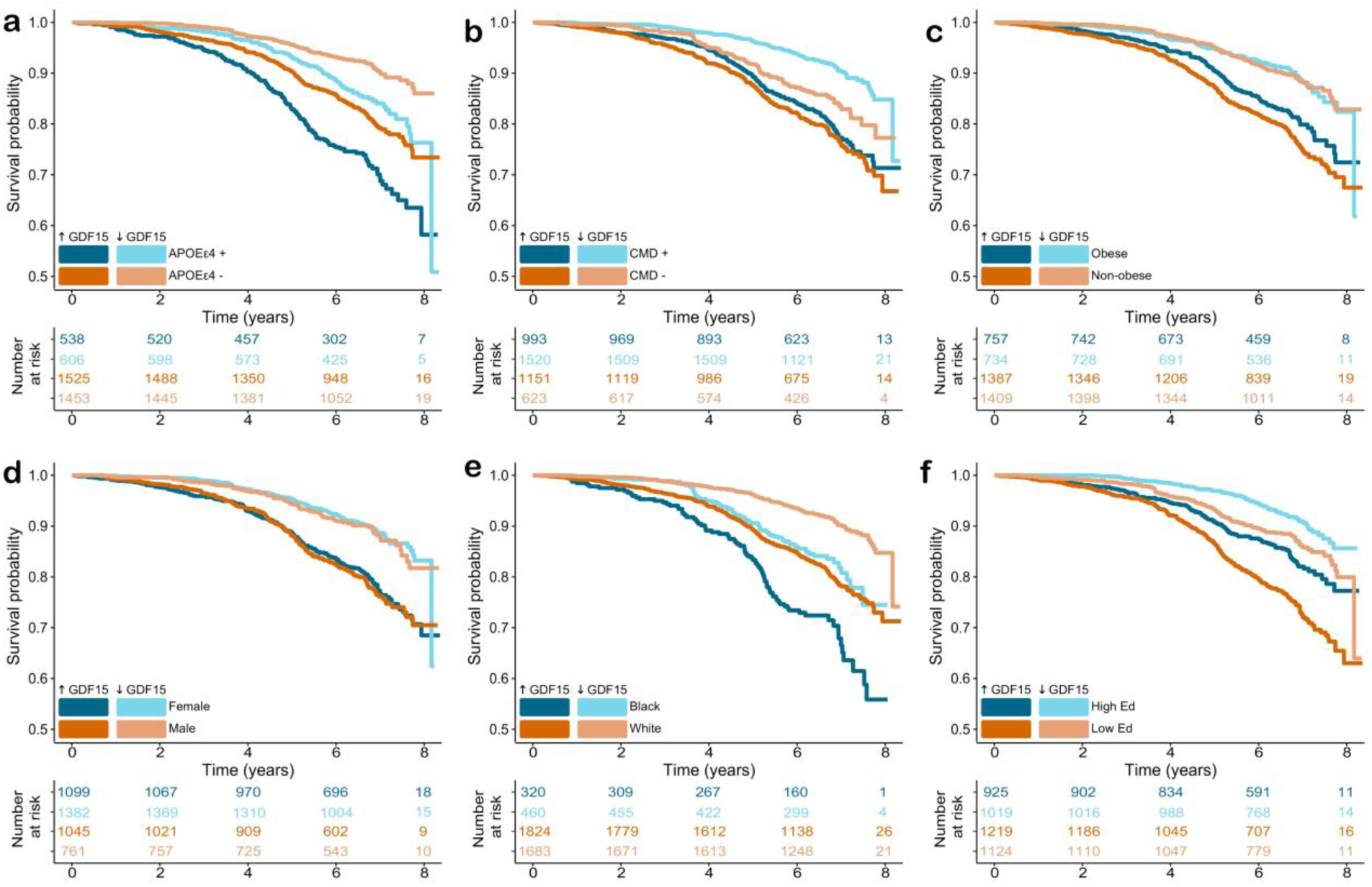
Kaplan-Meier curves showing plasma GDF15’s relationship to the probability of remaining free of ACD across late-life (7-year) in the Atherosclerosis Risk in Communities (ARIC) study. Associations of plasma GDF15 (high/low; median split) with late-life (7-year) ACD risk stratified by **a.** APOE*ε*4 status, **b.** cardiometabolic disease, **c.** obesity, **d.** sex, **e.** race, and **f.** educational attainment. *Abbreviations*: ACD; all-cause dementia; CMD, cardiometabolic disease; Ed, education.

**Extended Data Figure 3.**
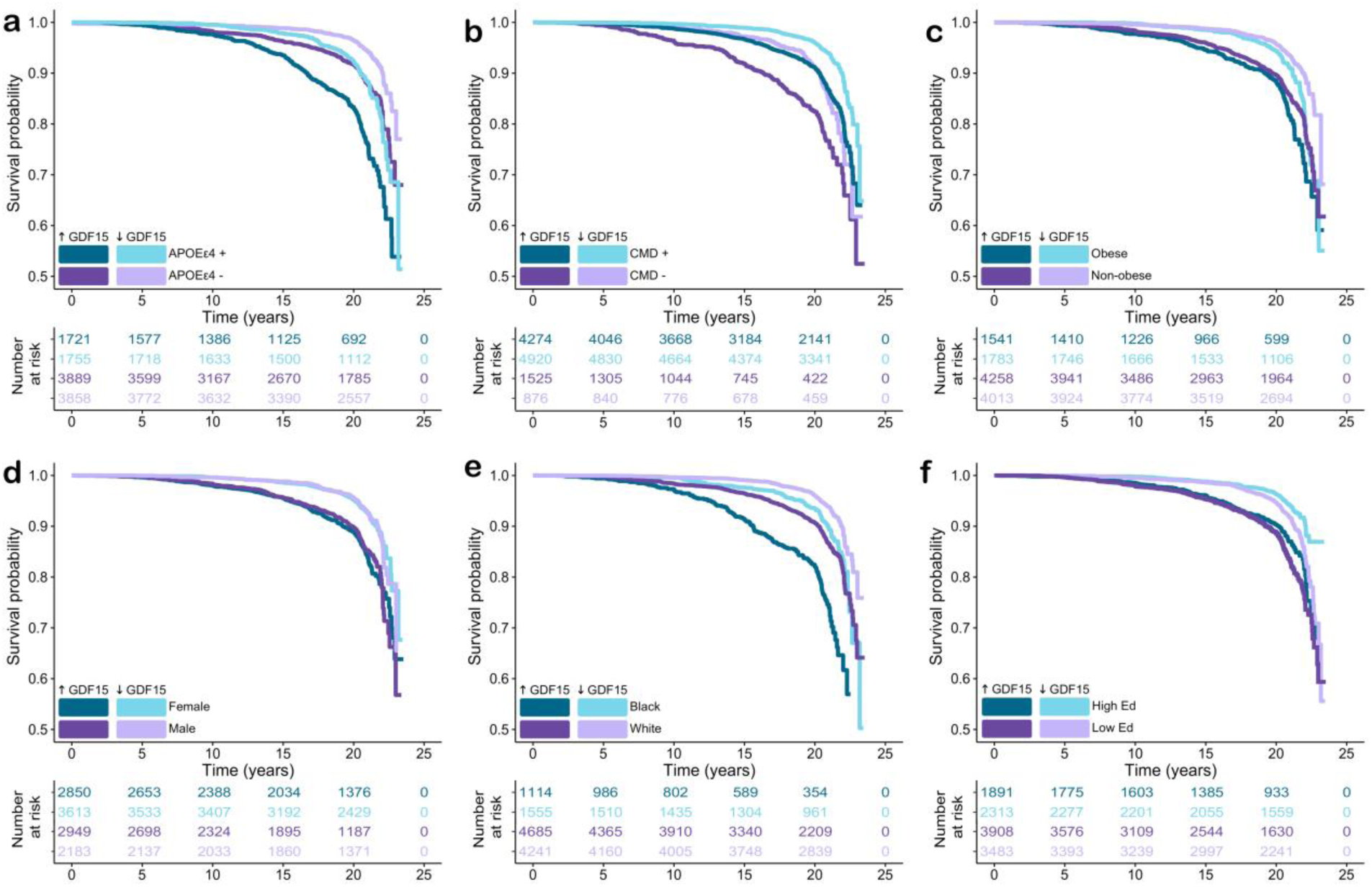
Kaplan-Meier curves showing plasma GDF15’s relationship to the probability of remaining free of ACD across midlife (20-year) in the Atherosclerosis Risk in Communities (ARIC) study. Associations of plasma GDF15 (high/low; median split) with midlife (20-year) ACD risk stratified by **a.** APOE*ε*4 status, **b.** cardiometabolic disease, **c.** obesity, **d.** sex, **e.** race, and **f.** educational attainment. *Abbreviations*: ACD, all-cause dementia; CMD, cardiometabolic disease; Ed, education.

**Extended Data Figure 4.**
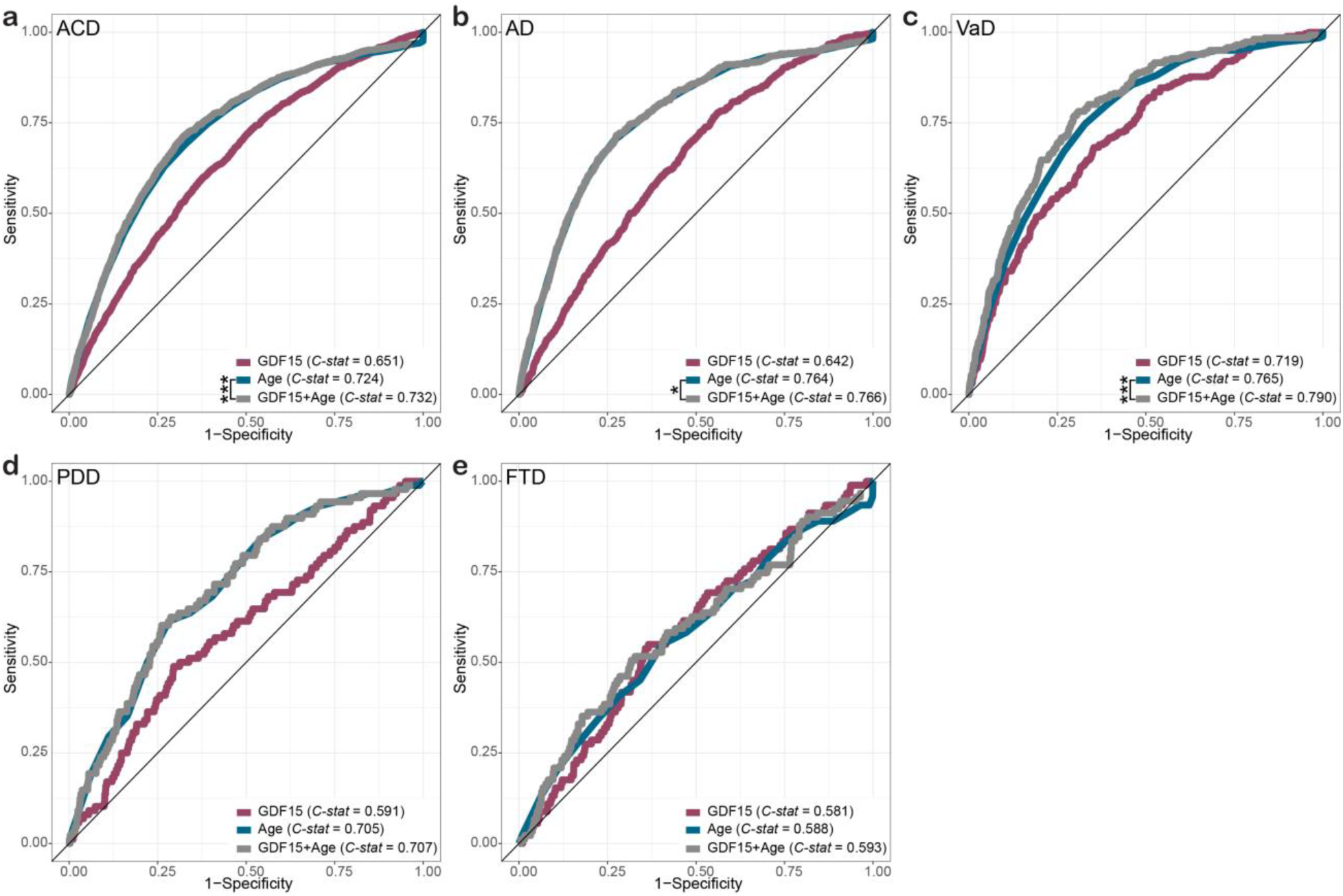
Predictive performance of plasma GDF15 for dementia risk by etiology in the UK Biobank (14-year follow-up) ROC curves representing the classification of 14-year incident dementia status by GDF15 levels alone, age alone, and GDF15 combined with age according to dementia subtypes, including **a.** all-cause dementia (ACD), **b.** Alzheimer’s disease (AD), **c.** vascular dementia (VaD), **d.** Parkinson’s disease dementia (PDD), and **e.** frontotemporal dementia (FTD). p: ***<0.001; *<0.05

**Extended Data Figure 5.**
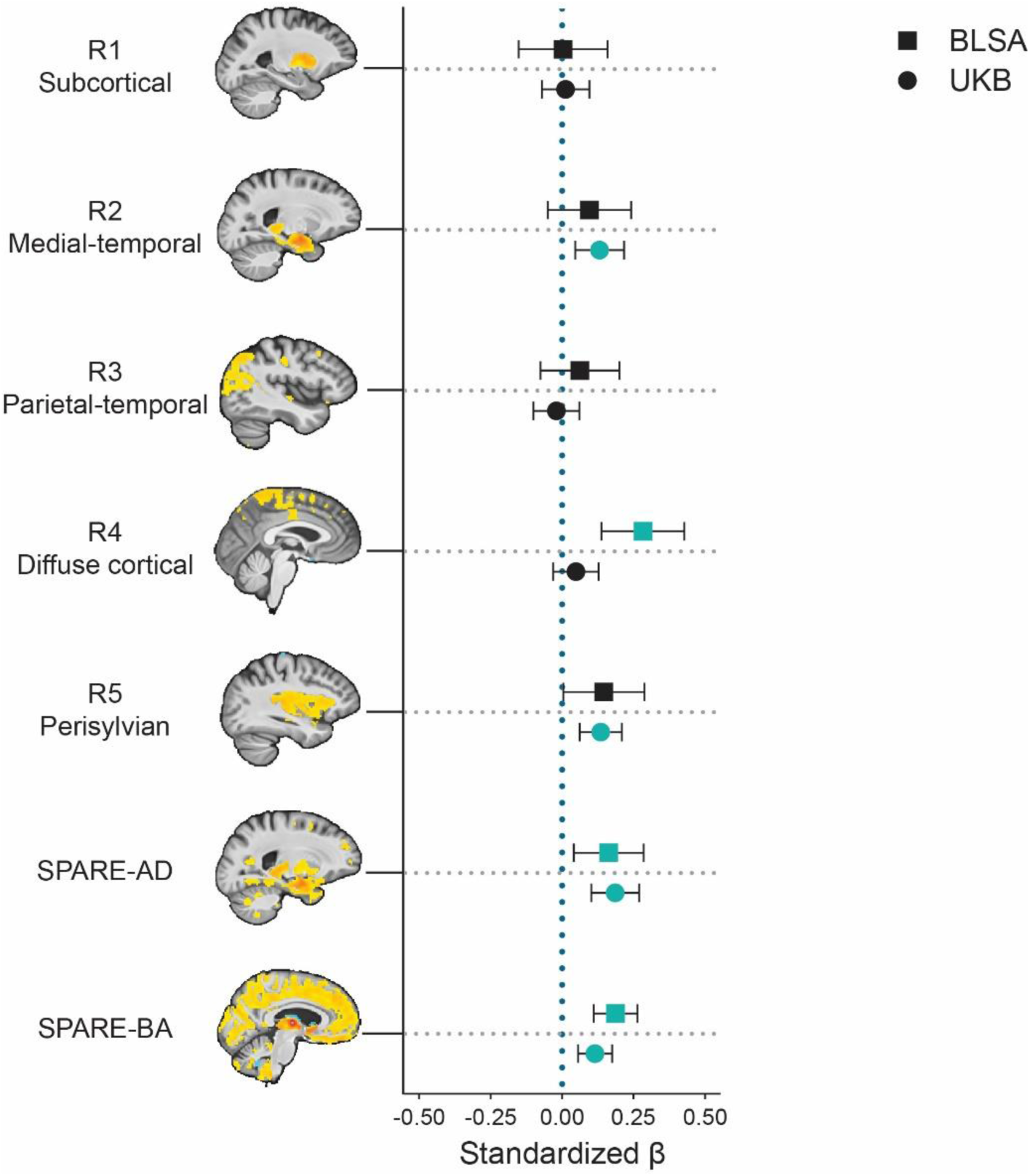
GDF15 relationship with machine-learning derived patterns of brain atrophy in the BLSA and UKB SPARE-AD and SPARE-BA were computed with support vector machine models to distinguish cognitively normal from clinically diagnosed AD individuals and to estimate the biological age of a participant’s brain structure, respectively. R index measures were computed with a semi-supervised deep representation learning approach (Surreal-GAN) to capture heterogenous brain volume differences between younger (<50 years old) and older (>50 years old) adults. Images on the left y-axis are representative voxel-wise images from BLSA participants.

**Extended Data Figure 6.**
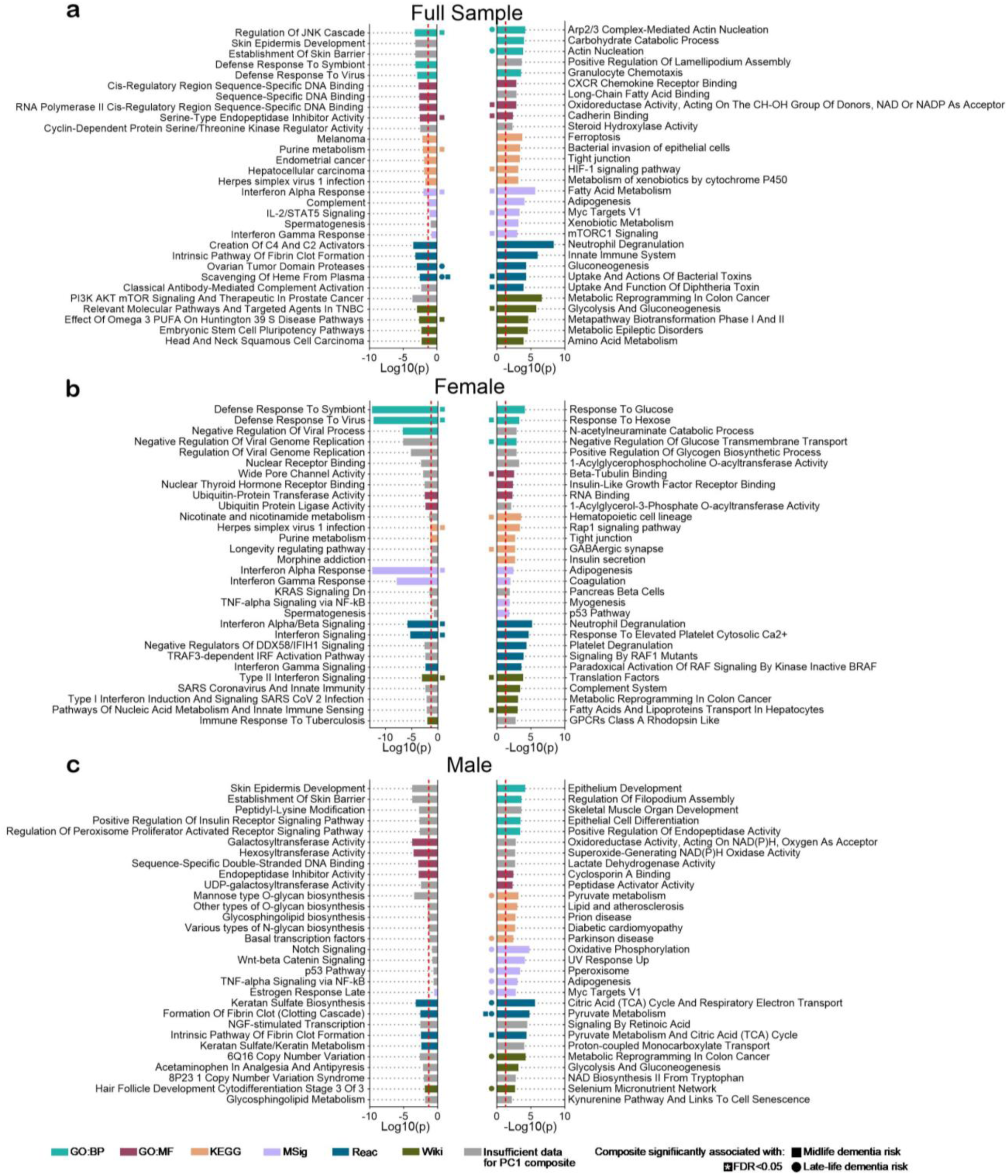
Pathways enriched following treatment with rhGDF15 compared to controls Proteins significantly affected by rhGDF15 treatment were analyzed for pathway enrichment using Enrichr^90–92^ in (a) the full sample of cell donors (N = 6), (b) female donors only (N = 3), and (c) male donors only (N = 3). For each comparison, the top five most annotated biological pathways were identified separately for upregulated and downregulated proteins. Gray bars indicate pathways for which no overlapping proteins were detected between the two measurement modalities—cell lysate mass spectrometry and ARIC participant plasma—preventing the calculation of principal component scores for those pathways. For additional annotation, see **Supplementary Table 20**.

